# Nucleotide motif-guided selection of plasma microRNA biomarkers for organ injury prediction in trauma

**DOI:** 10.1101/2025.03.02.25323184

**Authors:** Boyang Ren, Ruoxing Li, Chien-Yu Lin, Chanhee Park, Sheng Wang, Andrew O. Suen, John Kessler, Shiming Yang, Rosemary Kozar, Lin Zou, Brittney Williams, Ziyi Li, Peter Hu, Wei Chao

## Abstract

**BACKGROUND:** Trauma remains a leading cause of morbidity and mortality in part due to secondary organ injury and infection. Yet, our ability to predict the downstream pathophysiologic responses leading to organ injury and adverse outcomes is limited. Extracellular microRNAs (ex-miRNAs) as a DAMP can drive innate immune response and organ injury. Here, we tested plasma miRNAs as predictive biomarkers for organ injury in trauma.

**METHODS:** Twelve miRNAs were selected based on RNAseq and pro-inflammatory nucleotide motifs identified by machine learning. Digital PCR was employed to quantify plasma miRNAs and Luminex to measure trauma injury markers. Multivariate Random Forest models were built to assess the predictive performance of the miRNA biomarkers.

**RESULTS:** We identified a set of five nucleotide motifs that can predict the pro-inflammatory property of plasma miRNAs with a sensitivity of 84% and specificity of 69%. There was a marked and severity-dependent increase in the plasma miRNA biomarkers and numerous trauma injury markers at time of admission. The plasma concentrations of these miRNA biomarkers were highly correlated with the injury markers linked to various trauma endotypes. AUROC analyses indicated that the miRNA biomarkers possess strong diagnostic abilities and prediction in overall severity, organ injury, metabolic acidosis, coagulopathy, and innate inflammation in the trauma (n=48) but not sepsis (n=47) cohort. In a combined cohort, miR-224-5p and miR-145-5p exhibited a superior performance in differential diagnosis of trauma and sepsis with AUROC of 0.90 and 0.91, respectively.

**CONCLUSION:** The panel of plasma miRNAs are specific biomarkers with strong diagnostic and prognostic performance in trauma-induced organ injury.

## INTRODUCTION

Trauma is accountable for 13% of death and remains the leading cause of mortality among individuals age 1-44 years in the US.^1^ While advancements in the clinical management of traumatic injury has improved survival, the morbidity and mortality of severe injury is still unacceptably high in part due to secondary multi-organ injury and infection.^2^ Current treatment of trauma remains limited to supportive therapies and targeting individual pathophysiology. Moreover, trauma patients are heterogeneous in demographics, causes of injury, underlying pathophysiological responses, and responses to treatments, and as a result, their post-injury clinical trajectories are often unpredictable. These challenges highlight the need for the tools that could help predict the critical pathophysiological changes as well as the downstream adverse clinical outcomes such as coagulopathy, endotheliopathy, organ injury, and death after initial trauma.

Trauma elicits rapid activation of innate immunity and repair mechanisms in an attempt to restrict the damage.^3–5^ However, the innate immune activation can become grossly imbalanced and drive excessive inflammatory response, endothelial injury, cellular barrier dysfunction, and coagulopathy when initial injury is severe and sustained.^3, 6^ In the absence of infection, danger-associated molecular patterns (DAMPs), released from injured cells, are the presumed driver in maintaining and exacerbating the sustained activation of the innate immune system via pattern recognition receptors and subsequent organ injury.^3, 6^ As such, a forward feedback loop of tissue damage is formed leading to inflammation that in turn causes further cellular and organ impairment.

Recent studies have found that extracellular RNA, derived from the plasma, tissues, and cells under various pathological conditions such as trauma and sepsis, is a strong innate immune activator.^7–10^ Plasma RNAseq revealed that extracellular microRNAs (ex-miRNAs) are the dominant biotype of plasma small RNA and are markedly differentially expressed in trauma patients, demonstrating a unique signature of plasma miRNAs in trauma.^10^ miRNAs are small, single-stranded (ss) noncoding RNA ^11^ that primarily regulate gene expression inside the cell by binding to target mRNAs, leading to mRNA degradation and translation inhibition.^12, 13^ However, increasing studies have shown that miRNAs also exist in extracellular spaces, including circulation^14–17^ and various body fluids,^18–20^ where some, not all, of them can function as a DAMP by activating innate immune response via Toll-like receptor 7 (TLR7)^7–9^. In trauma and sepsis, ex-mRNAs have been proposed to act as a key driver in the innate immune response via TLR7 signaling and the downstream organ injury.^10, 21, 22^ However, the potential of plasma ex-miRNAs as biomarkers to predict the downstream trauma pathophysiology and clinical outcome has not been investigated.

In this study, we develop a computer exhaustive search algorithm and identified multiple nucleotide motifs uniquely presented in pro-inflammatory, but not in non-inflammatory, miRNA sequences. The utility of the nucleotide motifs to identify and predict the proinflammatory property of ex-miRNAs is tested and validated in different sets of miRNAs. Based on the plasma miRNA profiles and guided by the novel nucleotide motifs, we select a panel of 12 plasma miRNAs as biomarker candidates for testing. In a cohort of 48 patients with polytrauma, we find that the number of this panel of miRNA biomarkers are markedly upregulated at time of admission, tightly correlated with their injury severity score (ISS) and a wide spectrum of trauma pathophysiological markers linked to organ injury, coagulopathy, endotheliopathy, and inflammation. Moreover, the plasma miRNA biomarkers exhibit a strong performance in predicting secondary organ injury markers and ISS in trauma and a superior ability to differentiate trauma and sepsis in a combined cohort.

## METHODS

### Human subjects

This prospective observational trauma cohort study was conducted at the Shock Trauma Center of the University of Maryland. The inclusion criteria include: 1) patients at age between 18-45 of male and female, 2) patients with serious injury defined as one of the following: blunt injury to chest, abdomen, or pelvis, two or more long-bone fractures (tibia, femur, or humerus), and traumatic amputation proximal to wrist or ankle. Exclusion criteria include: 1) vulnerable population (pregnancy and prisoners), 2) two or more systemic diseases such as hypertension, diabetes, and smoking, 3) penetrating injury to the chest or abdomen, 4) unknown time of injury, 5) suspected toxic ingestion, 6) non-survivable brain injury, CT evidence of intracranial hemorrhage, 7) burns > 50% BSA, and 8) immunosuppressed. For the healthy cohort, age and sex-matched between age 18-45 were recruited with the exclusion criteria of 1) vulnerable population and 2) subject with two or more systemic diseases. Informed consent was obtained for each subject or their legally authorized representative. Patients were treated according to standard of care per institutional and national guidelines. We divided the study cohort by allocating 70% of the first-recruited trauma patients (n=34) and age- and sex-matched healthy controls (n=17) to the Training set and the remaining 30% of trauma patients (n=14) and healthy controls (n=7) to the Validation set. Sepsis patients (n=47) admitted to the intensive care unit and the age- and sex-matched healthy cohorts (n=24) were recruited in a prospective study. These patients were subsequently diagnosed with sepsis based on Sequential Organ Failure Assessment (SOFA) scores and confirmed through blood culture results.

### Animal use

C57BL/6 mice were purchased from Jackson Laboratory. Animal procedures were approved by the Institutional Animal Care and Use Committee (IACUC) of University of Maryland, Baltimore. Mice were euthanized under general anesthesia and bone marrow cells were isolated from femurs and tibiae under sterile conditions as described previously.^10^

### Computer searching algorithm for miRNA pro-inflammatory motif identification

The algorithm consists of three main components: building a motif list from s training set of 33 miRNAs, selecting pro-inflammatory motifs, and executing an automated motif-finding algorithm to narrow down to the most essential motifs. 1) Motifs ranging from two to ten nucleotides long within miRNA sequences in the training dataset were generated, including those with wildcards. 2) Three exclusion criteria were applied to the motif list; First, motifs that appear in non-inflammatory miRNA sequences; Second, motifs that showed up in less than 25% of pro-inflammatory miRNAs in the training set were excluded. Third, duplicate motifs defined by the same core sequence with differently existed wildcards on either end of the core sequence will be assess for their fitness in the training set, if the performance of duplicates were the same, sequence with the shortest length will be kept. 3) Finally, an automated motif-finding algorithm to pinpoint the most relevant motifs linked to inflammatory responses were developed based on a penalized LASSO (least absolute shrinkage and selection operator) regression model. A 5-fold cross-validation was used to tune the hyperparameter, Lambda, that controls the strength of the L1 regularization term in the LASSO model was adjusted so that the model retained the highest F1 performance score while utilizing the minimal number of non-zero coefficient motifs. Detailed workflow for the motif algorithm can be found in Supplementary Table 1.

### Patient clinical data

For trauma patients, blood samples were obtained at time of arrival. De-identified patients’ demographic and clinical information was obtained for further analysis. The clinical information included patients’ demographics, injury mechanism and pattern. The clinical metrics included injury severity score (ISS), aspartate aminotransferase (AST), alanine transaminase (ALT), alkaline phosphatase (Alk phos), total bilirubin, blood urea nitrogen (BUN), creatinine (Cr), lactate, prothrombin time (PT), partial thromboplastin time (PTT), international normalized ratio (INR), and platelet count (PLT), white blood cell (WBC), hemoglobin (Hb), hematocrit (Hct), and potassium (K).

### Plasma processing

Whole blood samples were collected in K_2_-EDTA tube and centrifuged at 1000 *g* for 10 minutes at room temperature. Supernatants were centrifuged at 1000 x *g* for 10 minutes at 4°C, transferred, and centrifuged again at 14,000 x *g* for 10 minutes at 4°C. Plasma was stored at -80°C until further use.

### Plasma RNA extraction and miRNA quantification by digital PCR

Plasma RNA was extracted using Qiagen miRNeasy Serum/Plasma Advanced Kit. Specifically, after adding lysis buffer, 250 μL of human EDTA-plasma was mixed with the spiked-in cel-miR-39-3p and proceeded to RNA purification. Purified RNA was eluted in 20 μL nuclease-free water. Six μL of RNA was used for cDNA synthesis using Qiagen miRCURY LNA RT Kit. The LNA-enhanced primers were designed for Qiagen miRCURY LNA miRNA PCR Assays, and miRNA copy numbers were quantified by a digital PCR platform from Qiagen (QIAcuity One). To calculate the recovery rate and correct for the loss of miRNA during plasma RNA extraction, an equal quantity of the spike-in cel-miR-39-3p was used for cDNA synthesis, quantified using digital PCR without going through the RNA extraction step, and termed as the “added spike-in”. The plasma concentration of target miRNA (copies/mL) was calculated as: detected miRNA copies × normalizing factor (*added* spike-in / *detected* spike-in copies) / plasma volume used.

### miRNA mimics

Single-stranded miRNA mimics were synthesized as the same ribonucleotide sequence as human mature miRNAs with modification of phosphorothioate linkages in between the ribose to enhance nuclease resistance and cell transportation. MiRNA mimics were synthesized from Integrated DNA Technologies (IDT) and reconstituted in RNase-free H_2_O.

### Bone marrow-derived macrophages

Bone marrow-derived macrophages (BMDMs) were prepared from eight to 12-week-old male mice as described previously.^22^ Bone marrow cells from the tibias and femurs were suspended in RPMI 1640 medium supplemented with 10% fetal bovine serum, 5% horse serum,10 ng/ml M-CSF, and penicillin/streptomycin at a density of 2×10^6^/mL and cultured in 96-well plate at 100μL/well. Two days after incubation at 37 °C and 5% CO_2_, culture medium was changed once before miRNA treatment.

### Raw 264.7 macrophage cell line

Murine macrophage cell line Raw 264.7 were cultured in high glucose Dulbecco’s Modified Eagle Medium (DMEM) supplied with L-glutamine,10% and penicillin/streptomycin at a density of 3×10^5^/mL and cultured in 96-well plate at 100μL/well.

### miRNA treatment of cells

Cells were incubated for 2 hours with serum-free medium supplemented with 0.05% BSA. Synthetic miRNA mimics were packaged with Lipofectamine 3000 before added to cell cultures. For Test set 1 and Test set 2, miRNAs were titrated at the concentrations between 3 - 500 nM in cultured media. Eighteen hours later, media were assayed for CXCL2 production using ELISA. miRNAs that induced 1) a dose-dependent increase of CXCL2 in culture medium and 2) at lease 1.5-fold increase CXCL2 production were defined as pro-inflammatory miRNAs.

### Luminex multiplex for plasma injury marker quantification

The concentrations of plasma protein analytes were determined by Luminex Assay (R&D systems), a fluorescent bead-based multiplex immunoassay platform. The analyte panel specifically designed for this study included S100B (S100 calcium-binding protein B), enolase2, ANG1 (angiopoietin1), ANG2 (angiopoietin2), P-selectin, tissue factor, d-dimer, IL-6 (interleukin-6), TNFα (tumor necrosis factor alpha), IL-8 (interleukin-8), CXCL2 (Chemokine (C-X-C motif) ligand 2), syndecan1, vWF-A2 (von Willebrand factor A2 domain) and VCAM1 (vascular cell adhesion protein 1). Plasma samples were thawed on ice and centrifuged at 12,000 x *g* for 10 min. The supernatants were applied to the antibody-conjugated magnetic bead and proceeded further following the manufacturer’s instructions. The fluorescent antibody reaction was measured on a Luminex 200 machine from Luminex xMAP Technology and the analysis was performed with the Luminex xPONENT software.

### Predictive modeling and performance evaluation

The injury markers from the Luminex assays or clinical data were binarized based on the corresponding median values. Random forest modeling of all 12 miRNA biomarker candidates to each marker was built. Data were subjected to 2-fold cross-validation (CV) for 100 times before feeding to the model. The mean values of sensitivity and specificity were used for plot ROC curve and the reported 95% confidence intervals (95% CI) were calculated based on all AUROC results from the CV. Receiver operating characteristics (ROC) curve was used to evaluate the performance of the model, and area under ROC (AUROC) was calculated by the trapezoidal rule in R package pROC (v1.18.5).

### Data cleaning

Luminex data under detection levels were set to 0. Missing clinical data were noted N/A and skipped in plotting and analysis.

### Software and packages

R Statistical Software (v4.4.0), package pROC (v1.18.5), ROCR (v1.0-11), randomForest (v4.7-1.1), dplyr (v1.0.10), ggplot2 (v3.5.1), ComplexHeatmap (v2.20.0) and ComplexUpset (v1.3.3), Prism GraphPad 10 were used for statistical analysis and data plotting. Python (v3.10) was used for motif discovery.

### Power calculation

Based on our plasma RNA-Seq data from a small cohort of severely injured patients (N=10) and age and sex matched healthy controls (N=10),^10^ the statistical power reached 0.86 when miRNA biomarkers were measured in the cohorts of 48 trauma patients and 24 controls assuming standard deviation of 12% and a minimum difference in means of 30% between groups.

### Statistics analysis

Analysis of the data was performed using R Statistical Software (v4.4.0). Data were tested for normality with QQ plot and Shapiro-wilk test. For variable with a normal distribution (Shapiro-Wilk test, p > 0.05) in all comparison groups, a t-test with Benjamini-Hochberg correction was used. For variable that did not follow a normal distribution, a Wilcoxon test with Holm-Bonferroni correction was used. Spearman’s rank correlation coefficient (r) was used to assess the relationship between miRNA and measured variables. Statistical details of experiments can be found in the figure legends. The null hypothesis was rejected for *P* < 0.05.

### Study approval

The protocol for trauma human study was approved and conducted in accordance with the policies and procedures approved by the Institutional Review Board of the University of Maryland (IRB protocol: HP-0011660). The animal experimental protocols were approved by the Institutional Animal Care and Use Committee, University of Maryland School of Medicine (IACUC protocol: 0722002).

### Data availability

RNA-seq dataset were used for selection of test set miRNAs and are available through the Gene Expression Omnibus. Test set 1: GSE133733, Small RNA-Seq of mouse plasma from model of polymicrobial sepsis. Test set 2: GSE223151, Small RNA-Seq of human plasma samples and mouse polytrauma. MiRNA biomarkers candidates were selected based on GSE223151. All data reported in the study will be shared by the lead contact upon request.

## RESULTS

### Study design

**Fig. 1** illustrates the overall design of the current study – motif discovery, biomarker selection, and biomarker evaluation. Previous studies show that some, but not all, upregulated plasma miRNAs in mice and humans after traumatic injury promote cytokine and complement production via TLR7.^10^ In this study, we tested the hypothesis that specific nucleotide sequences or motifs exist in these small RNA molecules that determine the proinflammatory property of miRNAs and can be used to guide the selection of miRNA biomarker candidates for trauma-induced inflammation and organ injuries. To achieve this, we developed a computer exhaustive search algorithm and identified five nucleotide motifs embedded specifically in pro-inflammatory miRNAs (**Fig. 1, *left panel***). Next, to select miRNA biomarker candidates, 10 patients with severe injury and 10 healthy controls were analyzed for RNAseq-based plasma RNA profiles. Based on the differential expression (DE) of the plasma miRNAs and the presence of any one of the five nucleotide motifs in their structures, we selected a panel of plasma miRNA biomarker candidates that were upregulated in trauma patients and possessed a proinflammatory property (**Fig. 1, *middle panel***). Finally, we tested the panel of miRNA biomarker candidates in a cohort of 48 trauma patients and 24 healthy subjects for their association with various trauma pathophysiology markers known to be linked with organ injury, coagulation, inflammation, and endothelial activation. We also tested the performance of the miRNA biomarkers in predicting secondary organ injury after initial trauma (**Fig. 1, *right panel***).

**Figure 1.**
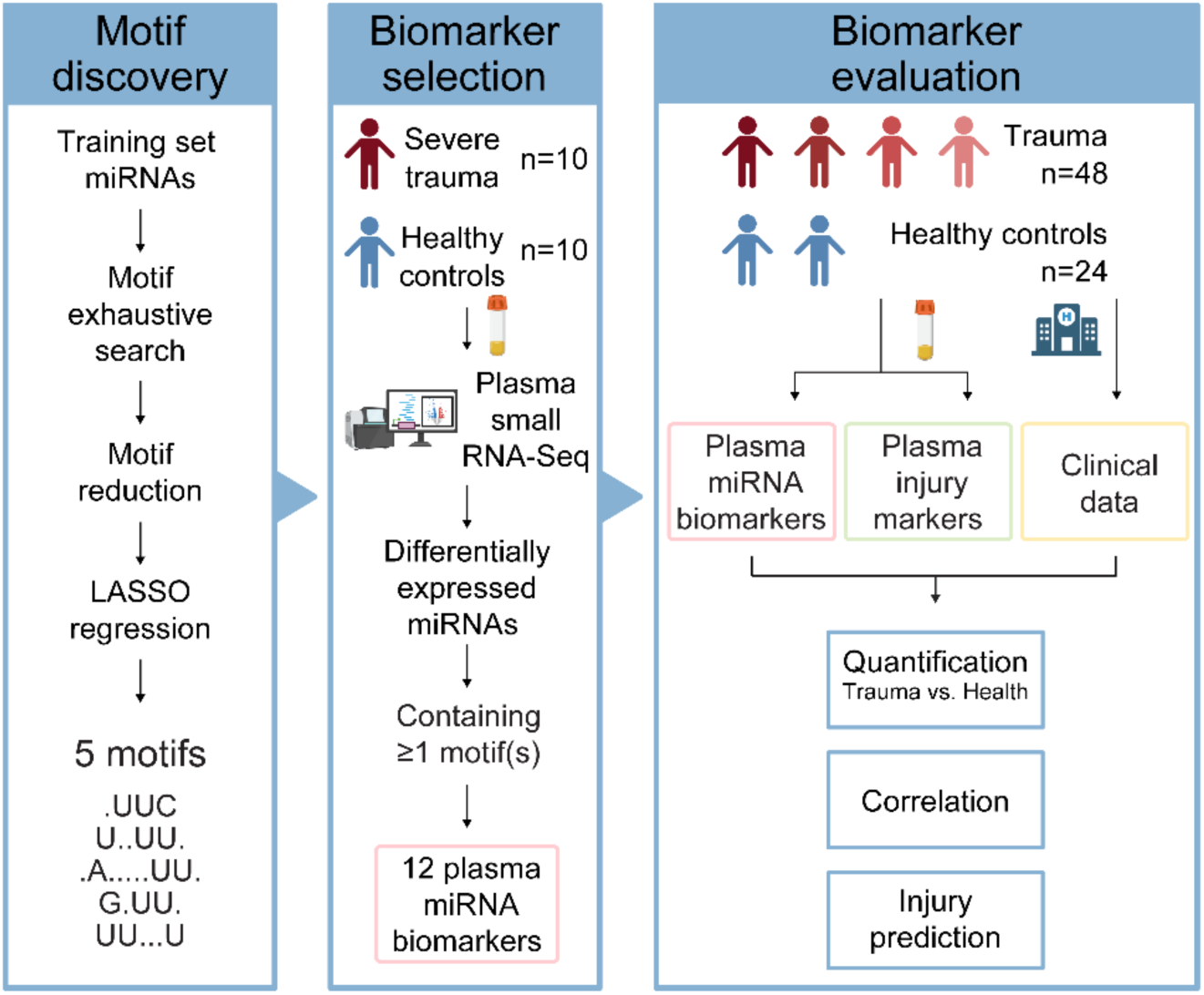
Overall study flow. The study consists of the following three parts: 1) Motif discovery (*left panel*): Five pro-inflammatory motifs are identified via a computer exhaustive search algorithm; 2) Biomarker selection (*middle panel*): A panel of 12 plasma miRNA biomarkers candidates are selected based on the DE of plasma miRNAs in trauma patients and guided by the pro-inflammatory miRNA motif; 3) Biomarker evaluation (*right panel*): The plasma miRNA biomarker candidates are quantified in the cohorts of control subjects (n=24) and trauma patients (n=48), tested for their correlations with various trauma pathophysiological markers, and evaluated for their predictive performance in secondary organ injury after trauma.

### Human study subjects

**Table 1** lists the demographic and clinical information of the trauma and healthy cohorts. The median age of the trauma and control groups was 29.0 and 30.5 years old, respectively, although the trauma group had a wider range of age distribution (**Fig. S1A**). Males represent 65% of the trauma cohort and 42% of the healthy group (**Fig. S1B**). The average ISS score was 15 (4-43) among the trauma cohort. Eighteen patients had severe injury with ISS > 15, an indicator of severe injury, and consequently had a longer hospital stay than those with less severe injury (**Fig. S1C-D**). **Table 1** and **Fig. S1E** illustrates the injury patterns present on arrival. Notably, while most of the patients had bone fracture(s), 18 patients also sustained at least one internal organ injury including lung, spleen, aorta, bowel, mesentery, kidney, and liver.

**Table 1.** Demographic and clinical information of human study subjects.

|  | **Control (n = 24)** | **Trauma (n = 48)** |
| --- | --- | --- |
| **Age, median (range)** | 30.5 (23-41) | 29.0 (19-53) |
| **Sex, male** | 10 (42%) | 31 (65%) |
| **Race** |  |  |
| White | 17 (71%) | 31 (65%) |
| African-American or Black | 3 (13%) | 13 (27%) |
| Asian | 1 (4%) | 1 (2%) |
| Hispanic/Latino | 2 (8%) | 0 (0%) |
| Other | 1 (4%) | 3 (6%) |
| **Mechanism of injury** |  |  |
| MVC | - | 30 (63%) |
| MCC | - | 4 (8%) |
| Ped MVA | - | 7 (15%) |
| Mechanical force | - | 3 (6%) |
| Fall | - | 4 (8%) |
| **Injury severity score (ISS)** | - | 15.23(4-43) |
| **Injury characteristics** |  |  |
| Bone fracture | - | 45 |
| Muscle injury | - | 10 |
| Lung injury | - | 8 |
| Splenic injury | - | 5 |
| Liver injury | - | 1 |
| Stomach injury | - | 1 |
| Bowel injury | - | 4 |
| Mesenteric injury | - | 2 |
| Renal injury | - | 1 |
| Aortic injury | - | 4 |
| **Laboratory** |  |  |
| INR | - | 1.033 (0.9-1.6) |
| PTT, s | - | 26.33 (21-85) |
| Platelets, $\times 10^3/\mu\text{l}$ | - | 257.2 (47-458) |
| Lactate, mmol/L | - | 3.165 (0.8-10.3) |

### Machine learning-guided discovery of pro-inflammatory motifs in miRNAs

To identify the nucleotide motifs embedded specifically in pro-inflammatory, but not in non-inflammatory, miRNAs, we developed a computer exhaustive search algorithm and took a machine learning approach for automated motif selection. As illustrated in **Fig. 2A**, we first generated a training data set containing 33 mouse miRNAs that are upregulated in the plasma of trauma mice.^10^ Among the 33 miRNA mimics tested in an in vitro cell system (cultured bone marrow-derived macrophages, BMDMs), 18 of them triggered a dose-dependent CXCL2 (Chemokine (C-X-C motif) ligand 2) production while the other 15 did not (non-inflammatory) as shown in **Fig. 2B**. Utilizing the search algorithm (**Table S1**), a list of 513,599 possible motifs ranging between 2mer – 10mer within the training set of 33 miRNAs were identified. The list of motifs was then shortened to 306 after we excluded those motifs that appeared in the non-inflammatory miRNAs, those that appeared in less than 25% of pro-inflammatory miRNAs, and those with duplicates. Thus, the remaining motifs were only present in the pro-inflammatory miRNAs (**Fig. 2A**). Next, we developed an automated motif-finding algorithm to pinpoint the most essential motifs linked to the pro-inflammatory property of miRNAs by utilizing the Least Absolute Shrinkage and Selection Operator (LASSO) regression model.^23^ By applying cross-validation and tuning the value of the regulator in the LASSO model, only essential motifs would be left with non-zero coefficients while retaining highest model performance monitored by F1 score (**Fig. S2**). As a result, five motifs – 1) .UUC, 2) U..UU., 3) .A. UU., 4) G.UU. and 5) UU U (“.” represents a wild card) – were identified specifically link to the 18 pro-inflammatory miRNAs of the training set (**Fig. 2A**), and a total of 41 pro-inflammatory motifs were discovered in the training set of miRNAs with a relaxed LASSO algorithm (**Table S2**). These data indicate that the five motifs are the most prevalent in the 18 pro-inflammatory miRNAs but completely absent in the 15 non-inflammatory miRNAs. **Fig. 2B** details the 33 miRNAs – 18 pro-inflammatory and 15 non-inflammatory – and their tested vs. motif-based predicted results of CXCL2 production in BMDMs. As illustrated in **Fig. 2B** and **Table 2**, the sensitivity of the individual motifs to identify pro-inflammatory miRNAs in the training set was between 56% ∼ 61% with the specificity at 100%. However, the five motifs together – *i.e.,* presence of any of the five – could distinguish pro-inflammatory miRNAs from non-inflammatory miRNAs with 100% sensitivity and 100% specificity.

**Figure 2.**
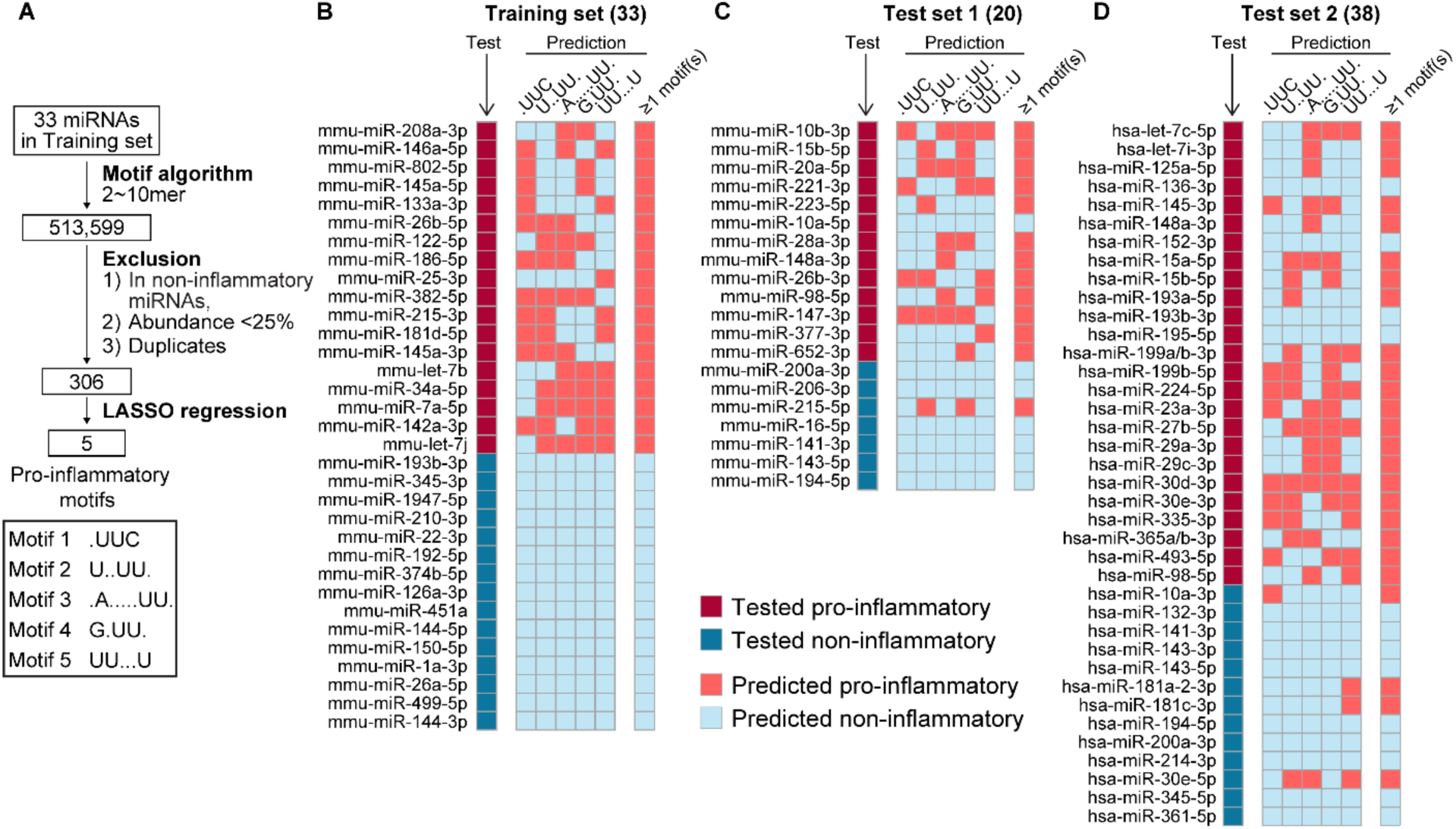
Machine learning-guided discovery of pro-inflammatory motifs in miRNAs. **A.** The motif identification algorithm and five essential motifs to predict pro-inflammatory miRNA. **B.** Training set – mouse miRNAs (n=33). *First column:* Test results of a dose-dependent CXCL2 response in BMDMs for each miRNA; *Middle 5 columns:* Prediction of pro-inflammatory miRNA by individual motif; *Last column:* prediction of pro-inflammatory miRNA by 5 motifs together. **C.** Test set 1 – mouse miRNAs (n=20). **D**. Test set 2 – human miRNAs (n=38).

**Table 2.**
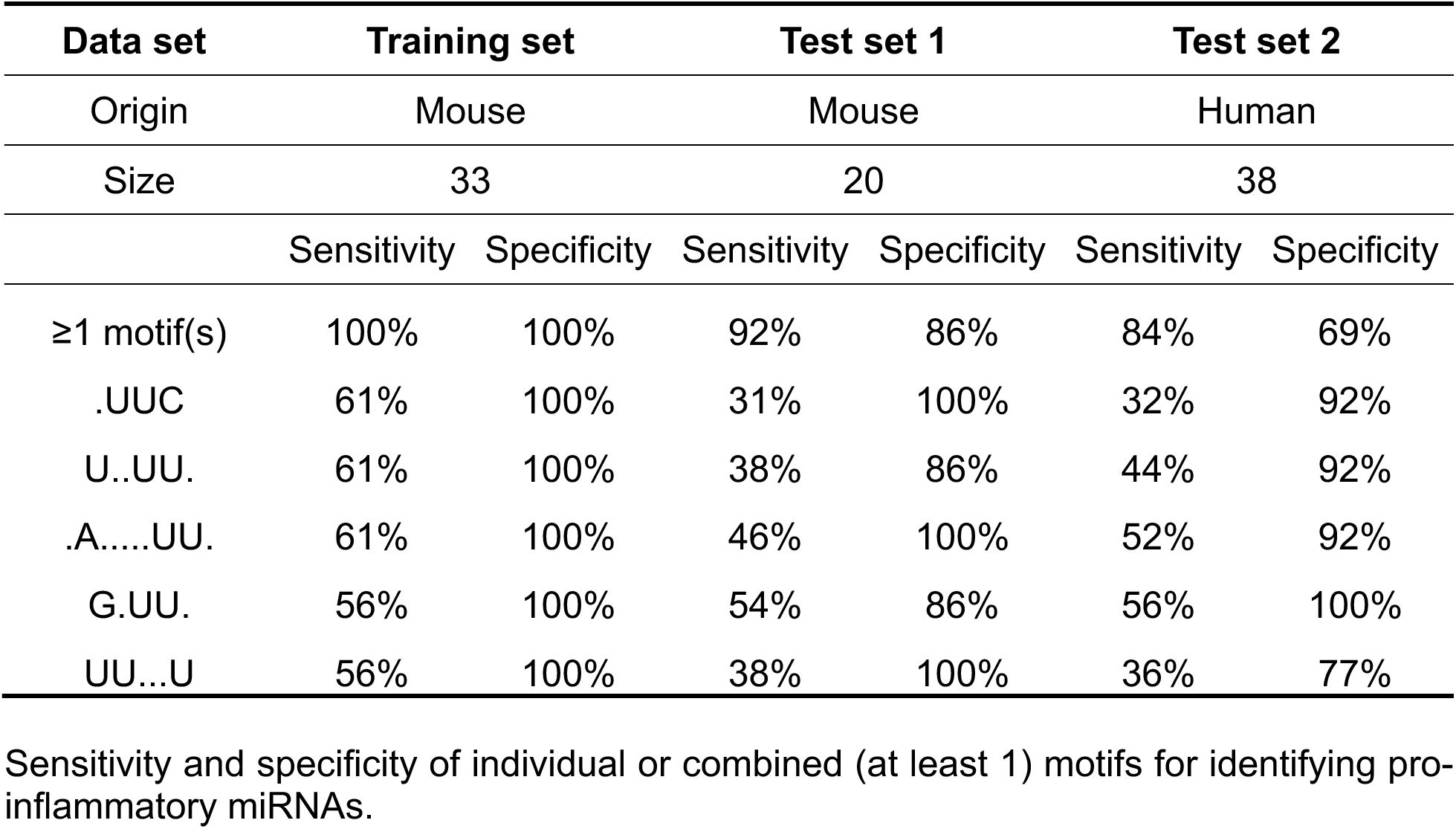
Performance of motifs in capturing pro-inflammatory miRNAs.

We next validated our algorithm in two separate sets of miRNAs - Test set 1 consisted of 20 mouse miRNAs upregulated in either a mouse model of sepsis or polytrauma in a previously published plasma small RNA-seq database (**Fig. 2C**).^10, 22^ Test set 2 consisted of 38 human miRNAs that were found upregulated in the plasma of our human trauma cohort based on previous plasma small RNA-Seq (**Fig. 2D**).^10^ Both sets of miRNAs were completely different from those in the training set. In these test sets, a dose-dependent cytokine CXCL2 production by cultured BMDMs treated with the miRNA mimics was considered the “ground truth” to define the “pro-” *vs* “non” inflammatory” miRNAs (**Fig. S3**). As illustrated in **Fig. 2C-D** and detailed in **Table 2**, the 5 motifs together (*i.e.,* any one of the five motifs) offered an overall prediction with 92% sensitivity and 86% specificity in Test set 1 and 84% sensitivity and 69% specificity in Test set 2. These data suggest that five miRNA nucleotide motifs possess strong sensitivity and reasonable specificity to predict the pro-inflammatory property of miRNAs and were used to guide the selection of miRNA biomarker candidates in trauma patients.

### Nucleotide motif-guided selection of proinflammatory miRNA biomarkers

To select plasma miRNA biomarker candidates, we first identified the upregulated plasma miRNAs in response to trauma based on our published RNA-Seq data (n=10, trauma vs n=10, health) at the time of admission.^10, 21^ Among 401 miRNAs detected in the human plasma, 151 were upregulated with a fold-change (FC) > 1.5 in the trauma patients. We then selected the most upregulated and abundant miRNAs in trauma plasma by the top 50% FC and top 50% count per million (CPM), applied the motif-finding algorithm to the top 38 miRNAs, and identified a panel of 12 miRNAs with at least one pro-inflammatory motif (**Table 3**).

**Table 3.**
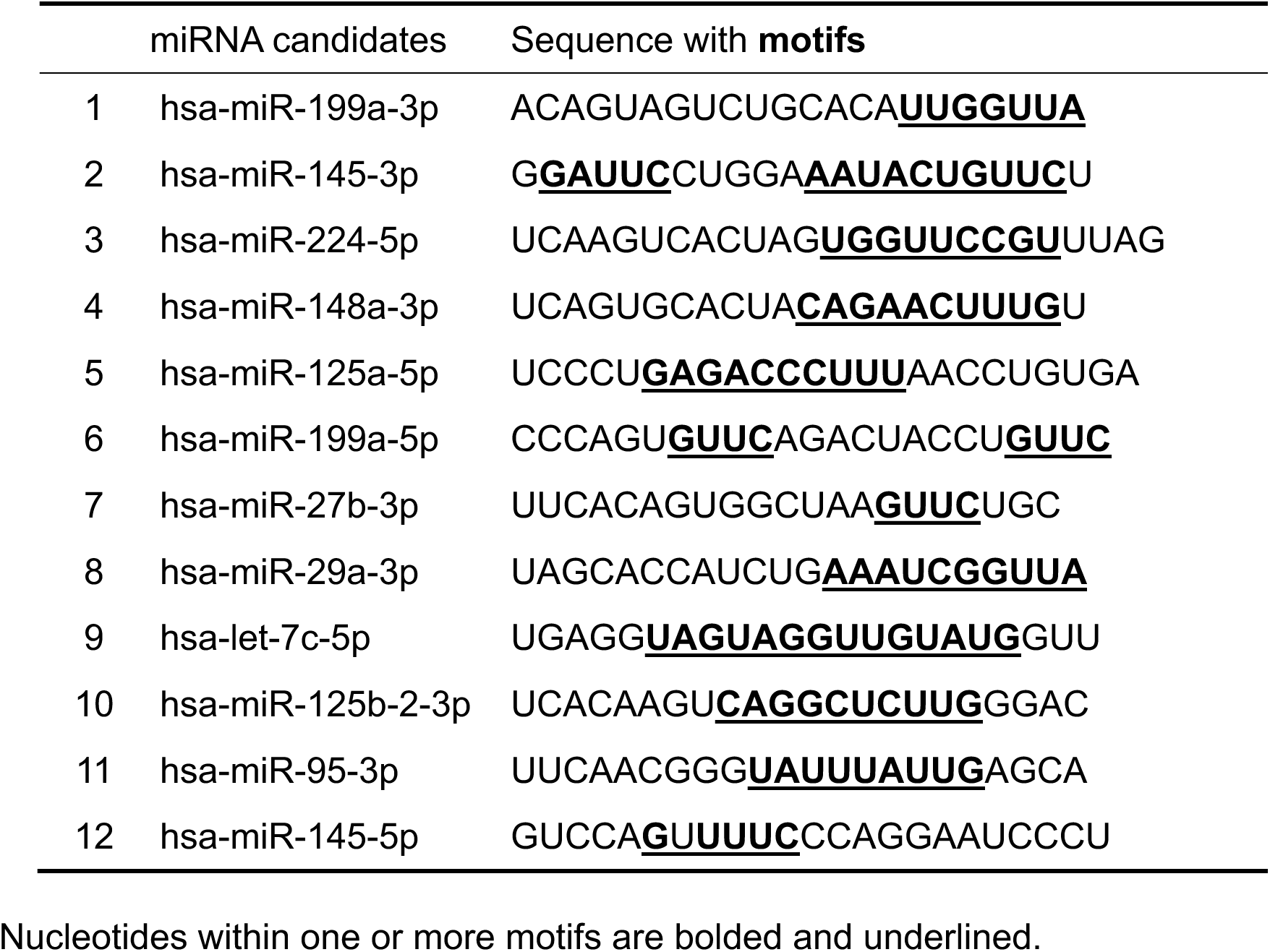
Motif-guided selection of miRNA biomarker candidates in trauma.

### Plasma miRNA biomarkers are markedly elevated in patients with blunt polytrauma

Next, we measured the plasma cell-free miRNA biomarker candidates used dPCR in the expanded cohorts of trauma (n=48) and healthy controls (n=24). We were able to quantify the copy number of the miRNA biomarkers and found that all 12 miRNA candidates were highly expressed and markedly upregulated in trauma patients as compared to the healthy controls (**Fig. 3A**). The fold-changes of trauma vs. control subjects ranged from 6.7 (let-7c-5p) to 78.2 (hsa-miR-145-5p). The fold-change was injury severity-dependent. In severely injured patients (ISS>15), 10 out of the 12 miRNA biomarker candidates exhibited significantly higher plasma levels than those in moderately injured patients (ISS≤15) (**Fig. 3B)**. The heatmap in **Fig. 3C** illustrates the three groups of cohorts – healthy control, trauma patients with ISS≤15, and trauma patients with ISS>15 – and their relative plasma expression of the 12 miRNA biomarkers. The heatmap reveals that the trauma patients were highly heterogenous in their plasma levels of the 12 miRNA candidates, but some trauma patients, particularly many severely injured with ISS>15, showed a cross-board upregulation of the 12 miRNA biomarkers.

**Figure 3.**
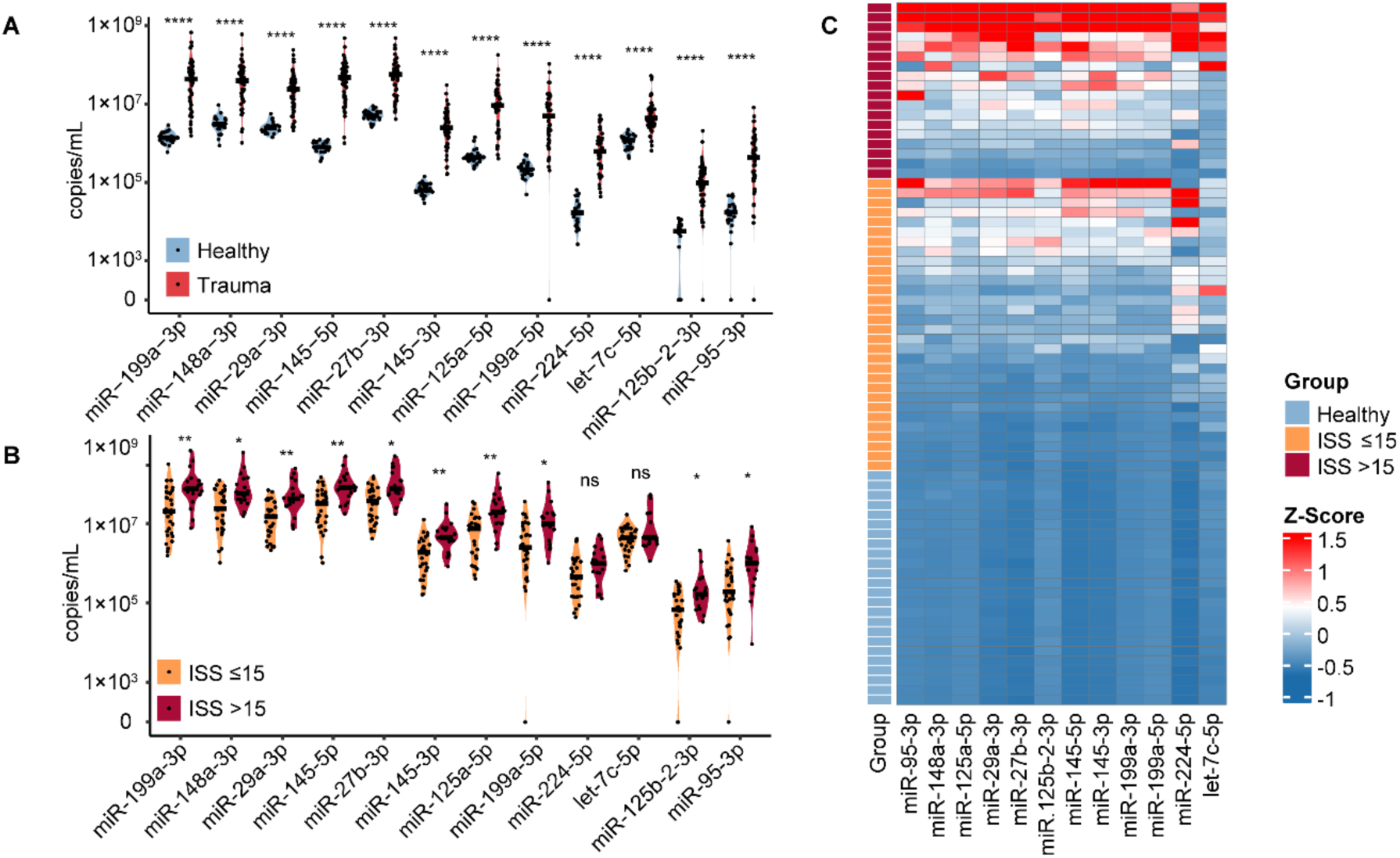
Plasma miRNA biomarkers are markedly elevated in patients with blunt polytrauma. **A.** Expression of plasma miRNA biomarkers in trauma patients versus healthy controls. Undetectable values were shown as zeros on y-axis. (Healthy: n=24, Trauma: n=48). **B.** Expression of plasma miRNA biomarkers between severe (ISS≤15: n=30) and moderate (ISS>15: n=18) trauma patients. **C.** Heatmap of plasma miRNA biomarker expression. Columns were clustered by Spearman’s correlation distance. Rows were grouped by injury severity (health, ISS≤15 and ISS>15) and within groups ranked by sum of Z-scores in descending order. Z score was calculated by subtracting the population mean from an individual miRNA copy number and then dividing the difference by the population standard deviation (n=72). Significance tested by Wilcox test with Holm-Bonferroni correction, ****, *p*<0.0001; ***, *p*<0.001; **, *p*<0.01; *: *p*<0.05. Each horizontal bar in A-B represents a median value.

### Plasma miRNA biomarkers are closely correlated with trauma endotype markers

The pathophysiology of trauma-induced secondary injury is complex. Immediate effects of trauma include the direct and indirect mechanical force on tissues, which induces tissue damage, contusions, hemorrhage, fractures, and disrupted barrier immune defenses.^3, 5^ Subsequent innate immune responses driven by DAMPs lead to excessive inflammation, endothelial injury, and coagulopathy. These pathophysiological events, often interlinked, result in hemodynamic instability, microvascular thrombosis, vascular leak, tissue hypoxia, secondary organ injury, and other clinical complications. To capture these critical events, we designed an extensive panel of mediators and injury or endotype markers that are linked to the trauma pathophysiology. Utilizing a Luminex-based multiplex assay system, we measured 14 such endotype markers present in the plasma at the time of admission, which included 1) various organ injury markers, 2) pro-coagulants, 3) pro-inflammatory cytokines and chemokines, and 4) mediators of endothelial activation. As illustrated in **Fig. 4A**, there was a marked and injury severity-dependent increase in the brain injury markers S100B^24, 25^ and enolase^25^ among the trauma cohort. A similar increase was observed with the lung injury markers of ANG2/ANG1.^26^ D-dimer, a fibrin degradation product, was markedly elevated, so were the two plasma pro-coagulants P-selectin and tissue factor in these trauma patients. Traumatic injury also led to vascular inflammation as evidenced by a significant increase in the pro-inflammatory mediators IL-6, TNFα, IL-8, and CXCL2 in the plasma. Also observed was marked endothelial cell activation and injury. Syndecan-1, a proteoglycan expressed on endothelial cells and a marker for endothelial injury, was notably increased in severely-injured patients with ISS >15. Both vWF-A2 and VCAM-1 exhibited an injury severity-dependent and marked increase after trauma. We next performed Spearman’s correlation analyses between the panel of 12 miRNA biomarkers and the various plasma protein injury and inflammatory markers. As illustrated in **Fig. 4B**, we discovered that within the trauma cohort, majority of the 12 ex-miRNA biomarker panel was either moderately (*r* > 0.4, *p* < 0.01) or strongly (*r* > 0.7, *p* < 0.01) correlated with various plasma inflammatory and injury markers, such as brain injury (S100B and enolase 2), coagulation (P-selectin, tissue factor and D-dimers), inflammation (IL-6 and IL8), and endothelial injury (syndecan-1, vWF-A2). It is noteworthy that the injury markers like ANG-1, CXCL-2, and VCAM-1, which did not exhibit severity-dependent upregulation, also showed no correlation with the plasma miRNA biomarker levels.

**Figure 4.**
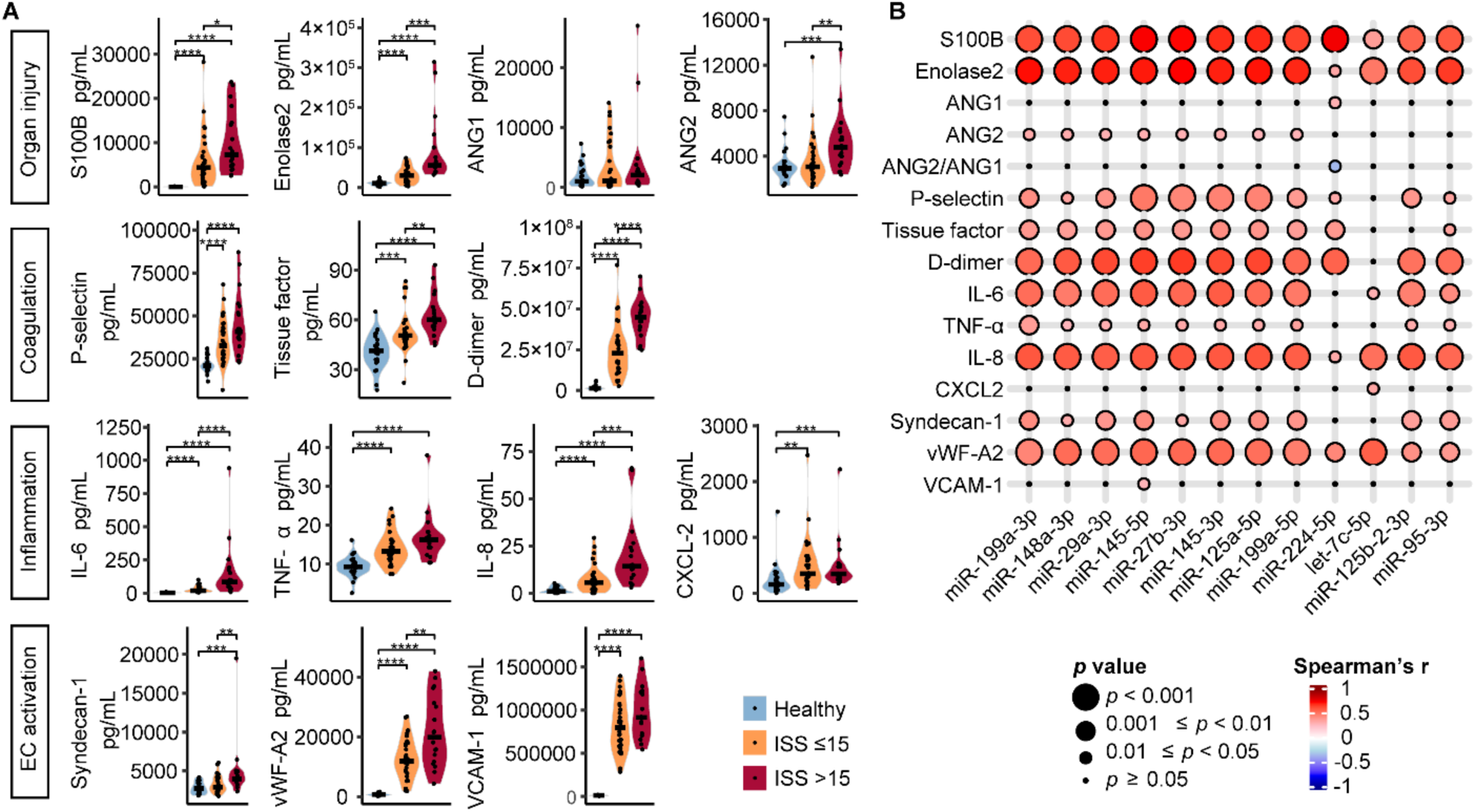
Plasma miRNAs biomarkers are closely correlated with trauma pathophysiology. **A.** Multiplex quantification of plasma mediators and organ injury markers. (Healthy controls: n=24, ISS≤15: n=30, ISS>15: n=18). Significance tested by t test with Benjamini-Hochberg correction (for normally distributed P-selectin, vWF-A2 and VCAM-1) or Wilcox test with Holm-Bonferroni correction (for the other mediators). ****, *p*<0.0001; ***, *p*<0.001; **, *p*<0.01; *, *p*<0.05. Each horizontal bar represents a median value. **B.** Correlation of miRNA biomarker candidates with protein mediator and organ injury markers within the trauma cohorts (n=48).

### Plasma miRNA biomarkers are correlated with AST/ALT, lactate, and ISS in trauma

Standard and routine clinical blood tests at the time of admission provide critical information about trauma pathophysiology such as organ injury, coagulopathy, and tissue metabolism. As shown in **Fig. 5A**, the cohort of trauma patients at the early stage exhibited only limited secondary organ injury with elevated AST and ALT, two liver injury markers, and serum lactate, a sensitive indicator of tissue ischemia and anaerobic metabolism. Most of the trauma patients only exhibited normal coagulation function as evidenced by normal PTT, PT, and INR, and normal kidney function with normal creatinine and BUN. While white blood cell counts were abnormally high in injured patients, platelets and hemoglobin remained normal. Clinical injury markers that remained within the normal range showed no correlation with plasma miRNA expression, whereas with the injury markers that were elevated at admission, the miRNA biomarkers exhibited a close correlation including AST/ALT, lactate, and somewhat with WBC (**Fig. 5B**). Importantly, with exception of miR-224-5p and miR-7c-5p which did not exhibit severity-dependent elevation (**Fig. 3B**), the miRNA biomarkers were closely correlated with the patients’ overall injury severity score (ISS), which was based on injury severity in 6 different body regions and usually calculated at the late stage when patients were discharged from the trauma center (**Fig. 5B**).

**Figure 5.**
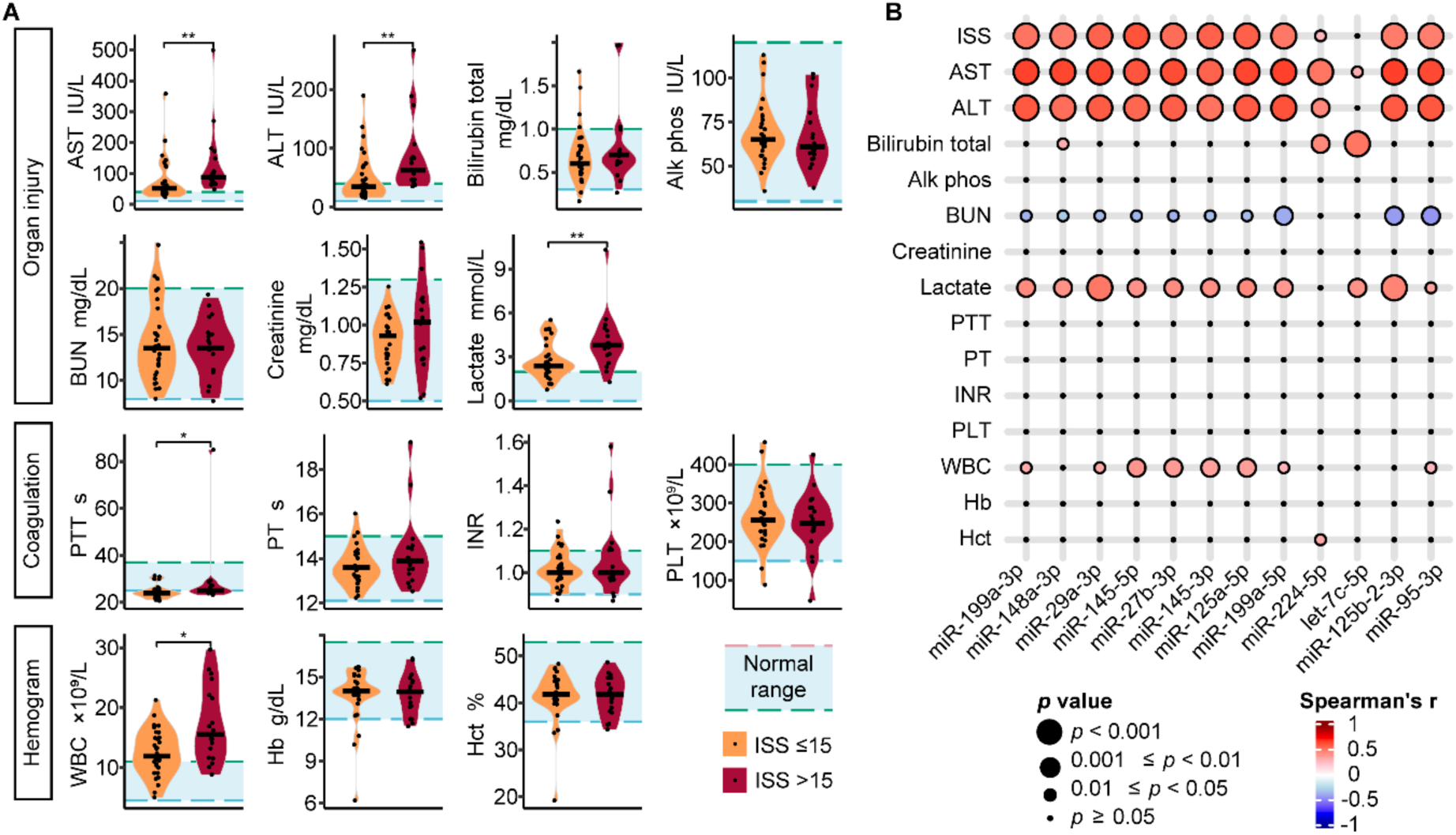
Plasma miRNA biomarkers are correlated with early organ injury in trauma. **A.** Clinical serology data at time of trauma center arrival. For ALT (alanine transaminase), Alk phos (alkaline phosphatase), BUN (blood urea nitrogen), Creatinine, Lactate, PLT (platelet count), WBC (white blood cell), Hb (Hemoglobin) and Hct (hematocrit), ISS≤15: n=30, ISS>15: n=18; for AST (aspartate transferase), Bilirubin total, INR (the international normalized ratio) and PT (prothrombin time), ISS≤15: n=29, ISS>15: n=17; for PTT (partial thromboplastin time), ISS≤15, n=27; ISS>15, n=16. Significance tested by *t* test with Benjamini-Hochberg correction (for normally distributed BUN, Creatinine, PLT and WBC) or Wilcox test with Holm-Bonferroni correction (for the other clinical data)., ****, *p*<0.0001; ***, *p*<0.001; **, *p*<0.01; *, *p*<0.05. Each horizontal bar represents a median value. **B.** Correlation of miRNA biomarker candidates with clinical injury markers within trauma cohorts. (For ISS (Injury severity score), ALT, Alk phos, BUN, Creatinine, Lactate, PLT, WBC, Hb and Hct, n=48; for AST, Bilirubin total, INR and PT, n=46; for PTT, n=43).

### Performance in prediction of trauma injury endotypes using miRNA biomarkers

In a cohort of 34 trauma and 17 healthy controls, the logistic regression model demonstrated that the panel of 12-miRNA biomarkers could distinguish the trauma and healthy groups with a cross validated Areas Under Receiver Operating Characteristics Curves (CV AUROC) of 0.98 [95% CI, 0.95-1] (**Fig. 6A**). When the upper 98% values of the endotype markers in the control group were used as the classifiers, the miRNA panel demonstrated a strong performance in identifying various trauma endotype markers in brain injury, coagulation, inflammation and EC activation with the respective CV AUROC values of 0.90 [0.80-0.98] (Enolase 2), 0.97 [0.95-1] (S100B), 0.85 [0.76-0.94] (P-selectin) and 0.86 [0.94-0.99] (D-dimer), 0.97 [0.93-1] (IL-6), 0.93 [0.86-0.98] (IL-8), and 0.96 [0.93-1] (VCAM-1) and 0.97 [0.93-1] (vWF-A2) (**Fig. 6A**, solid red line). The predictive performance of the models was further validated in a separate set of trauma (n=14) and healthy (n=7) cohorts where high CV AUROCs were also achieved (**Fig. 6A**, green dashed line). In comparison and as a negative control, demographic factors such as age and sex had a weaker performance with the CV AUROC between 0.55-0.67 (**Fig.6A**, gray line). Not surprisingly, for some endotype injury markers that were not increased or slightly elevated in the trauma cohort − such as the lung injury markers ANG1 and ANG2, tissue factor, TNFα, and syndecan-1 − the miRNA biomarker panel did not perform as well as the ones noted above (**Fig. S4A**).

**Figure 6.**
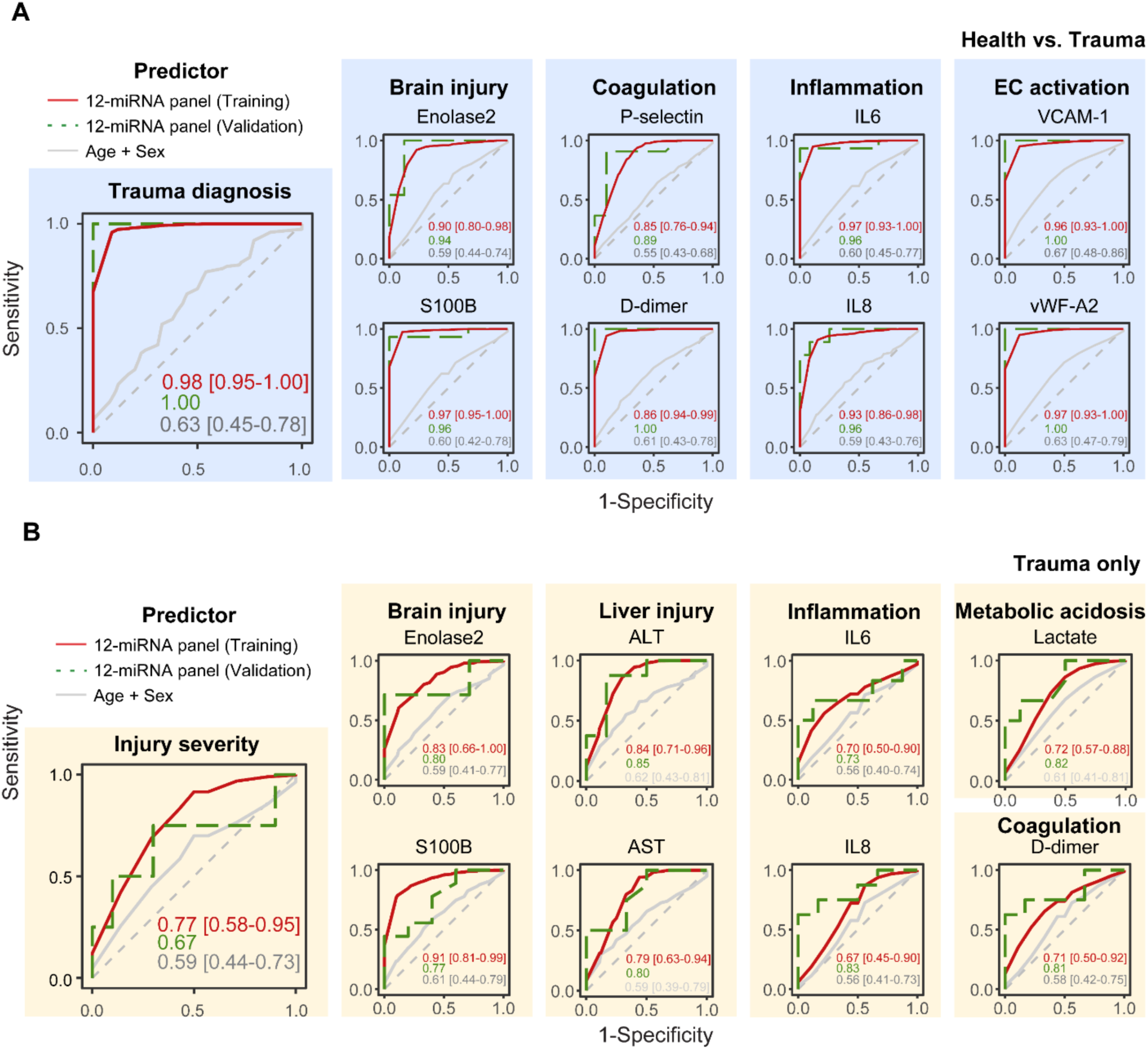
Diagnosis and multi-organ injury prediction by the miRNA panel. **A.** Receiver operating characteristic (ROC) curves for predicting trauma diagnosis and identifying abnormal levels, defined by exceeding normal range in health controls. **B.** ROC curves for predicting severe injury, defined by exceeding median level in the trauma patients except ISS, in which >15 was used. Prediction by 12-miRNA panel or Age+Sex in training set (N=51, 34 trauma and 17 healthy controls) and 12-miRNA panel in validation set (N=21, 14 trauma and 7 healthy controls) are shown. Random Forest regression was used for modeling within Training set, with 100 times 2-fold cross-validation. AUROC values were presented with 95% confidence intervals.

Furthermore, we evaluated the predictive performance of the 12-miRNA panel in distinguishing between severe and moderate injury. As illustrated in **Fig. 6B**, the CV AUROC values were 0.72 [0.55-0.88] for severe injury (based on their injury severity score (ISS) >15)^27^ and severe brain injury 0.83 [0.66-1] (Enolase 2) and 0.91 [0.81-0.99] (S100B). Similarly, the CV AUROC values for liver injury were 0.84 [0.71-0.96] (ALT), 0.79 [0.63-0.94] (AST), for inflammation, they were 0.70 [0.50-0.90] (IL-6) and 0.67 [0.45-0.90] (IL-8), For metabolic acidosis, it was 0.72 [0.57-0.88] (lactate), and for coagulation indicator D-dimer, it was 0.71 [0.50-0.92]. As anticipated, age and sex as a reference were not associated with the normality and severity of various organ injury and had near 0.5 AUROC values, indicating their poor performance in the trauma injury marker prediction. The severity-prediction model of the panel of miRNA biomarkers performed similarly in the validation set (**Fig**. **6B**, green dashed line). In comparison, the miRNA biomarker panel performed weak to distinguish the levels of coagulation factors (P-selectin, tissue factor, PTT, PT, INR), endothelial cell activation markers (syndecan-1 and VCAM-1), and lung injury markers (ANG1 and ANG2) within trauma cohorts largely due to the lack of either substantial post-trauma responses or severity-dependent increase (**Fig. S4B**). Finally, we evaluated whether ISS could distinguish the severity of the quantitative injury markers. The results indicated that ISS had a partial success in distinguishing brain injury severity (Enolase2, AUROC=0.70), plasma lactate (0.74), D-dimer (0.80) and IL6 (0.71) (**Fig. S5**).

### The miRNA biomarker panel exhibits disease-specific performance in trauma vs. sepsis

We tested the performance of the 12-miRNA panel of trauma biomarkers in separate trauma and sepsis cohorts. Our published plasma small RNAseq results^10, 21, 22^ revealed that among 212 upregulated miRNAs among the trauma and sepsis cohorts, only 51 (24.1%) of them were shared by both groups, whereas 100 (47.2%) were uniquely upregulated in the trauma cohort (**Fig. 7A**). Specifically, the 12-miRNA biomarkers were markedly upregulated in trauma patients but only modestly in septic patients (**Fig. 7B**). Digital PCR quantification of the plasma miRNAs in the trauma (trauma, n=48; control, n=24) and sepsis (sepsis, n=47; control, n=24) cohorts confirmed their preferential upregulation in the trauma cohort (**Fig. 7C**). Principle component analysis (**Fig. 7 D,E**) showed that 12-miRNA panel were partially separated between trauma and sepsis cohorts, whereas the commonly studies biomarkers – such as IL-6^28–31^, tissue factor^32^, and lactate^33^ – were completely shared between the trauma and sepsis patients. The major variance in patients were also better explained by 12-miRNA panel (PC1: 64.4%) than IL-6, tissue factor and lactate panel (PC1:45.0%). This indicated that the 12-panel of miRNAs better captured the difference between the two patient groups than IL-6, TF, and lactate, a group of well-known biomarkers of in both trauma and sepsis.

**Figure 7.**
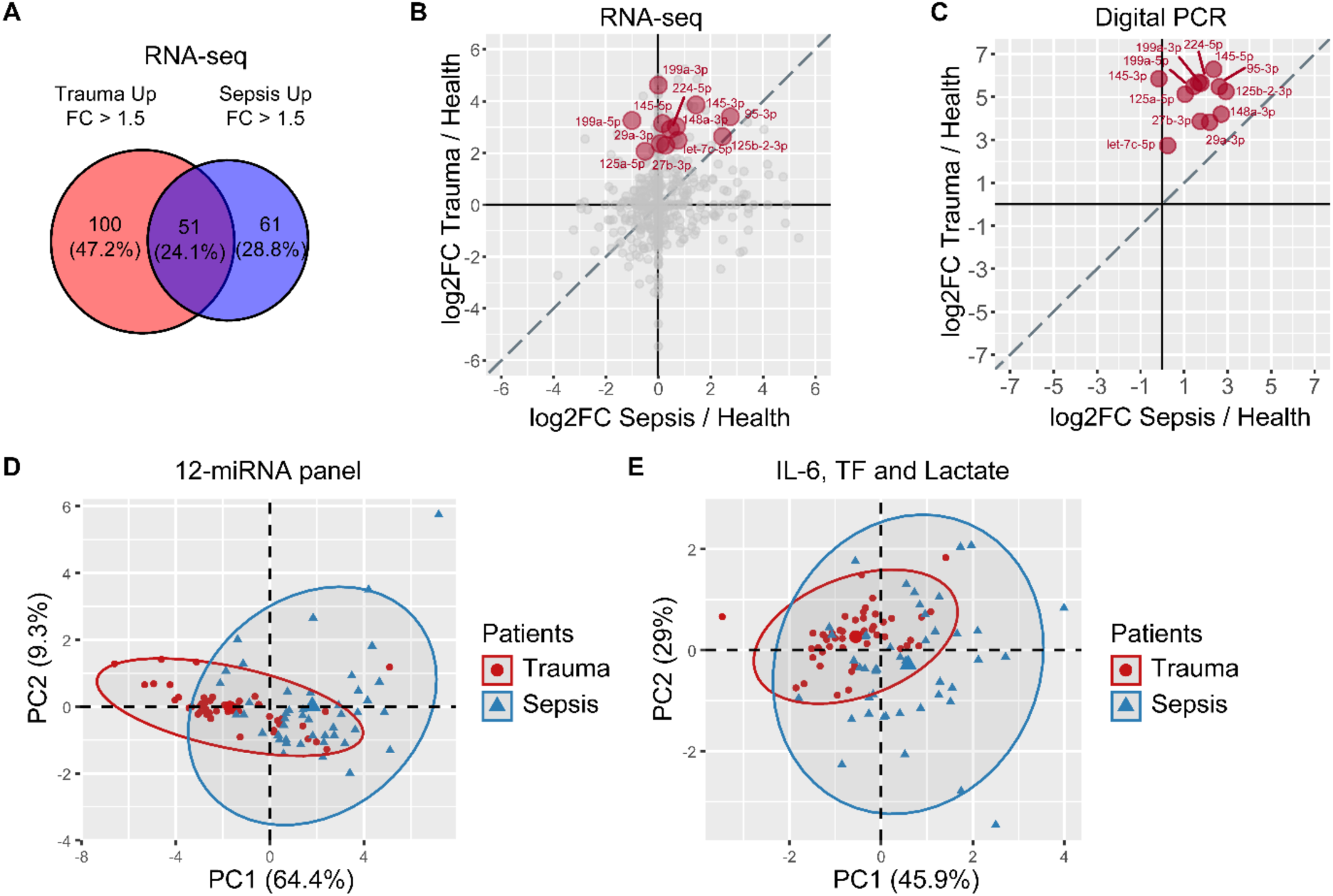
miRNA biomarkers exhibit preferential upregulation in trauma. **A.** Venn diagram showing overlapping of upregulated plasma miRNAs in trauma vs. sepsis cohorts based on RNAseq analysis (trauma, n=10; control, n=10). **B**. Fold change of the 12 trauma miRNA biomarkers in trauma (trauma, n=10; control, n=10) and sepsis (sepsis, n=10; health, n=8) cohorts. **C.** Fold change of the trauma miRNA biomarkers plasma concentrations as quantified by digital PCR in the trauma (trauma, n=48; control, n=24) and sepsis (sepsis, n=47; control, n=24) cohorts. **D.** Principal component analysis (PCA) for trauma and sepsis patients (n=95) by the level of 12 miRNA panel. **E.** PCA for trauma and sepsis patients (n=95) by the level of IL-6, tissue factor and lactate.

We then performed ROC analysis to evaluate the diagnostic performance of these biomarkers in the combined cohort of trauma and sepsis, and compared them with that of the known biomarkers IL-6, TF, and lactate.^34^ The selected miRNAs – such as miR-224-5p and miR145-5p – showed a strong performance with CV AUROC of 0.91 and 0.90 (**Fig. 8**) in differentiating trauma from septic patients. In comparison, IL-6, tissue factor, and lactate exhibited significantly lower CV AUROC values in their performance. The cut-off value of miR-224-5p for differential diagnosis of trauma vs. sepsis patients is 1.8×10^5^ copies/mL plasma, with sensitivity of 85% and specificity of 87%, whereas the cut-off value of miR-145-5p for the same diagnosis is 1.0×10^7^ copies/mL, with sensitivity and specificity both at 89%.

**Figure 8.**
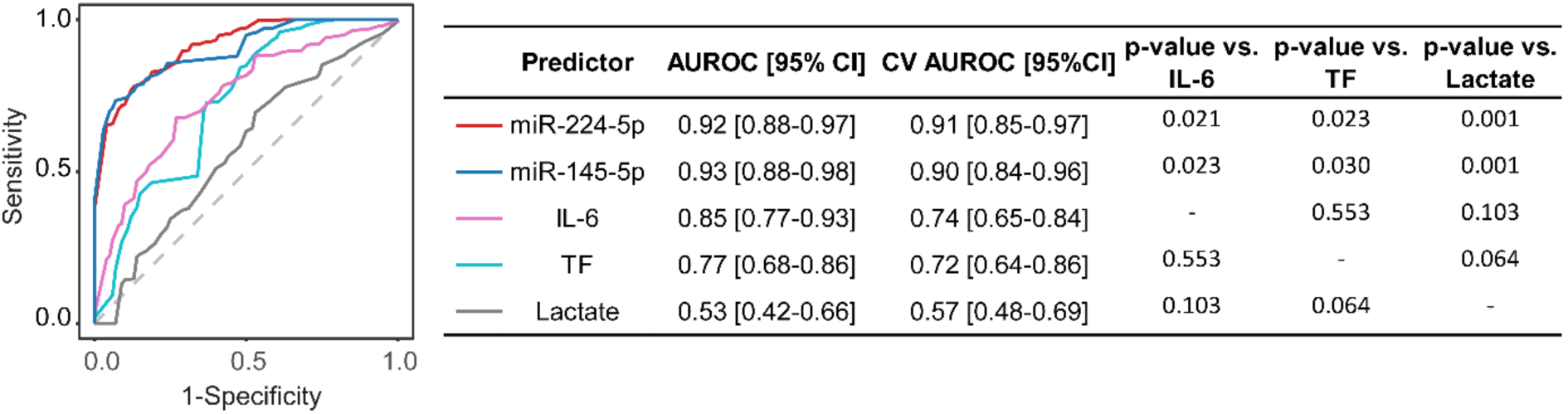
Comparison miRNAs and other known injury markers in the diagnostic performance in a cohort of trauma vs. septic patients. ROC curves of the Random Forest model for diagnosis of trauma versus sepsis (n=95) by individual predictors and their respective AUROC [95% CI], the cross-validated AUROC (CV AUROC) [95% CI] and the p values compared with IL-6, tissue factor (TF) and lactate as predictor. DeLong tests were performed for comparing two ROC curves.

Finally, we evaluated the predictive ability of miR-145-5p and miR-224-5p in the cohort of trauma vs. healthy controls. The CV AUROC of the two miRNAs in identifying trauma was 0.88 [0.77-1] and 0.98 [0.85 – 1.00] respectively, whereas their CV AUROC values were significantly lower in sepsis at 0.68 [0.63-0.90] and 0.56 [0.47 – 0.78], respectively (**Fig. S6A-B**). Similarly, for prediction of organ injury as measured by various injury markers of brain injury, coagulation, inflammation, and EC activation, miR-145-5p and miR224-5p performed consistently better in trauma with significantly higher CV AUROC values than in those of sepsis cohorts (**Fig. S6A-B**). In contrast, IL-6 showed similar predictive CV AUROC values in diagnosis and organ injury in trauma and sepsis cohorts (**Fig. S6C**). In comparison, tissue factor showed moderate to weak prediction performance in trauma or sepsis diagnosis and organ injury.

## DISCUSSION

Our findings provide proof-of-concept that plasma miRNAs could be used as biomarkers in traumatic injury that are not only closely associated with various signature markers linked to trauma pathogenesis such as inflammation, coagulopathy, and endotheliopathy, but also possess strong predictive abilities for these early pathological outcomes. Notably, these trauma endotype markers, while not standard clinical laboratory tests, offer a molecular lens into the wide spectrum of pathophysiological responses in the event of traumatic injury. These finding support the hypothesis that extracellular plasma miRNAs and the subsequent innate immune signaling are the molecular drivers that may be responsible for some of the key pathophysiological responses to traumatic injury such as inflammation and secondary organ injury.^10, 21^

Numerous trauma biomarkers have been reported, most of which are associated with downstream pathophysiology such as cell injury, endothelial activation, coagulopathy, and organ dysfunction^34–38^. For example, several cellular proteins released from injured cells, primarily neuronal cells, including Glial fibrillary acidic protein(GFAP), Ubiquitin C-terminal hydrolase-L1 (UCH-L1), S100B, and Neuron-specific enolase (NSE), are reported for diagnosing traumatic brain injuries.^39^ Other trauma biomarkers, such as IL-6, TNFα, C-reactive protein (CRP), and syndecan-1, are products of the downstream innate immune responses and endothelial injury to trauma and infection. As such, they are not disease-specific and often unable to distinguish between infectious and non-infectious systemic inflammatory response syndromes, such as sepsis and trauma. This limitation underscores the need for novel trauma biomarkers that are disease-specific and capable of predicting organ dysfunction and adverse outcomes with greater precision.

A number of studies have investigated the diagnostic and prognostic values of blood biomarkers, both proteins and nucleic acids, in traumatic brain injury (TBI) ^40–45^ and non-TBI polytrauma^46–49^. Several studies have shown that plasma miRNAs are differentially expressed upon TBI and some have been proposed as TBI biomarkers.^44, 50–52^ In our study, we selected the panel of 12 plasma miRNAs as biomarker candidates based on their large differential expression in trauma patients as compared with healthy subjects, their high abundance in the plasma samples, and the presence of pro-inflammatory motifs in their nucleotide sequences. The computer searching algorithm-based machine learning strategy enables us to identify and validate the five motifs that offer strong prediction for the pro-inflammatory property of the tested miRNAs with excellent sensitivity and moderate specificity. It is noteworthy that the five essential motifs all include a UU dimer, which may facilitate miRNA binding to TLR7 and thus enhance its pro-inflammatory property. A previous study demonstrates that successive uridine-containing single-stranded RNA fully or moderately bind TLR7, whereas single uridine-containing ssRNAs have reduced affinities.^53^ The importance of the UU dimer in one of the motifs, UU…U, has been evidenced by abolishment of miR-146a-5p-stimuated and TLR7-dependent inflammatory cytokine production when its UU…U motif was mutated to AU…U or UA…U.^22^ Thus, the identification of these specific motifs represents a unique strategy that links the inflammatory property of plasma miRNAs and miRNA biomarker selections in trauma and other clinical conditions where innate immune activation and inflammation are a critical part of disease pathogenesis, such as sepsis,^54^ shock,^3, 54^ reperfusion injury,^55, 56^ neurodegeneration,^57^ pain,^58^ and cancer metastasis.^59^

Our data demonstrated a disease-specific performance of the miRNA biomarker panel in trauma and sepsis diagnosis and injury prediction. RNAseq revealed distinct plasma miRNA profiles between the two criticaql conditions, with most of the 12 miRNAs showing greater upregulation in trauma as compared to sepsis, highlighting a trauma-specific miRNA signature. The PCA data suggest a better discriminatory power of the miRNA panel between trauma and sepsis when compared with the conventional biomarkers like IL-6, tissue factor, and lactate, which were completely overlapping between the two cohorts of patients. Moreover, two of the miRNA candidates, miR-224-5p and miR-145-5p, achieved high diagnostic accuracy in the combined cohort of trauma and sepsis, with AUROCs of 0.91 and 0.90, respectively, which significantly outperformed IL-6, tissue factor and lactate. These miRNAs exhibited trauma-specific predictive ability for diagnosis of organ injuries compared to sepsis, whereas IL-6 showed much similar performance in both conditions, reflecting its broad role as a pleiotropic inflammatory mediator commonly seen in trauma and sepsis. Enhanced specificity of biomarkers could enable more precise clinical diagnoses and targeted therapeutic strategies.

Our data revealed the diverse physiological and pathophysiological responses in the cohort of trauma patients such as liver injury, brain injury, coagulation activation, systemic inflammatory syndrome, and endothelial activation. It is noteworthy that there was no clinically recognizable coagulopathy or acute kidney injury in these patients given the normal INR and BUN/Creatinine at the time of admission. The majority of patients (63%) had relatively mild injury with ISS ≤ 15. The samples were collected very early at the time of arrival before severe clinical coagulopathy and acute kidney injury could potentially develop. Nevertheless, the finding that the plasma miRNA biomarkers exhibit a strong correlation with the individual protein mediator of injuries suggest a potential molecular link between the plasma miRNAs and the pathophysiological events. Our previous data have supported the role of extracellular miRNA and its downstream TLR7 signaling in inflammatory response,^22^ endothelial dysfunction,^60^ organ injury,^10, 21, 22, 60, 61^ and coagulopathy^62^ during sepsis and trauma.^21^ Together, these data support the hypothesis that these miRNA biomarkers are not only associated with the downstream pathophysiological changes in response to traumatic injury, but also likely play a contributory role.

Our data provide a proof-of-principle that the miRNA biomarker panel confers strong diagnostic and prognostic abilities in differentiating trauma from sepsis patients and predicting the severity of organ injury such as the liver and brain injury, overall trauma severity scores, metabolic acidosis, coagulation activation, and circulating cytokine storm. If validated in future prospective studies of larger independent cohorts, the plasma miRNA biomarkers could help to identify individuals who are at risk for secondary multi-organ injury and other clinical outcomes such as mortality following severe injury and as such, to guide individualized clinical management of trauma patients.

### Limitations of the study

Several limitations exist in the current study. While the ROC data in the training set was validated in a separate cohort, the sample size in both cohorts was small, and an external and larger trauma cohort will be needed to further test the utility of the miRNA biomarkers in order to generalize their clinical application. The small cohort size could impact the ROC analyses and performance assessment of the predictability of the miRNA biomarkers. Moreover, the cohort of blunt trauma patients in this study had relatively modest severity without associated mortality. While various trauma endotype mediators and injury markers were significantly increased following the initial trauma, most clinical organ injury markers remained within the normal ranges at time of trauma center arrival when blood samples were drawn. Analyzing longitudinal plasma samples in more severely injured patients in future prospective study will help quantify ongoing development of trauma-induced secondary organ injury and other adverse clinical outcomes, and further validate the prognostic value of these novel miRNA biomarkers.

## Data Availability

1. Data - All data reported in the study will be shared by the lead contact upon request
2. The code for statistical analysis and plotting in R program will be shared by the lead contact upon request.
3. Any additional information required to reanalyze the data reported in this paper is available from the lead contact upon request.

## ACKNOWLEDGEMENTS

This work was supported in part by the National Institutes of Health grants R35-GM140822 (WC) and R35-GM124775 (L.Z.). We thank Rashida Mohamed-Hinds from University of Maryland School of Medicine, for collecting and organizing the trauma clinical data. The graphic study design was created with BioRender.com.

## AUTHOR CONTRIBUTIONS

Conceptualization, P.H. and W.C.; Methodology, B.R., C-Y.L., R.L., C.P., S.Y.; Investigation, B.R., C-Y.L., R.L., C.P., Z.L., L.Z., B.W., P.H., and W.C.; Writing — original draft, B.R., C-Y. L., and W.C.; Writing — review & editing, Z.L., R.K., L.Z., B.W., P.H., and W.C.; Funding acquisition, W.C.; Resources, L.Z., B.W. and W.C.; Supervision, Z.L., P.H., and W.C.

## PATENT PENDING

The authors declare the following competing interests:

1. B.Y., S.Y., P.H., and W.C. are listed as inventors on a U.S. Provisional Application No. 63/734,468 entitled: “*Extracellular miRNA Motifs for Inflammation, Methods of Discovery and Uses Thereof*”
2. B.Y. and W.C. are listed as inventors on a U.S. Provisional Application No. 63/54,643 entitled: “*Methods for Determining Trauma-Induced Secondary Organ Injury*”.

## SUPPLEMENTAL INFORMATION

**Figure S1.** Demographic and clinical information of the control and trauma cohorts

**Figure S2.** Selection of essential motifs in LASSO model

**Figure S3.** Testing of miRNA mimics for CXCL2 production in cultured macrophages, related to Figure 2.

**Figure S4.** ROC analysis for injury prediction

**Figure S5**. Prediction of injury severity by ISS

**Figure S6.** Single predictor model for diagnosis and organ injury prediction.

**Table S1.** Computer exhaustive search algorithm

**Table S2.** Pro-inflammatory motifs identified with a relaxed LASSO algorithm

