## Supplemental Figures & Tables for "Nucleotide motif-guided selection of plasma microRNA biomarkers for organ injury prediction in trauma"

**SUPPLEMENTAL INFORMATION**

**Figure S1.** Demographic and clinical information of the control and trauma cohorts

**Figure S2.** Selection of essential motifs in LASSO model

**Figure S3.** Testing of miRNA mimics for CXCL2 production in cultured macrophages, related to Figure 2.

**Figure S4.** ROC analysis for injury prediction

**Figure S5**. Prediction of injury severity by ISS

**Figure S6.** Single predictor model for diagnosis and organ injury prediction.

**Table S1.** Computer exhaustive search algorithm

**Table S2.** Pro-inflammatory motifs identified with a relaxed LASSO algorithm

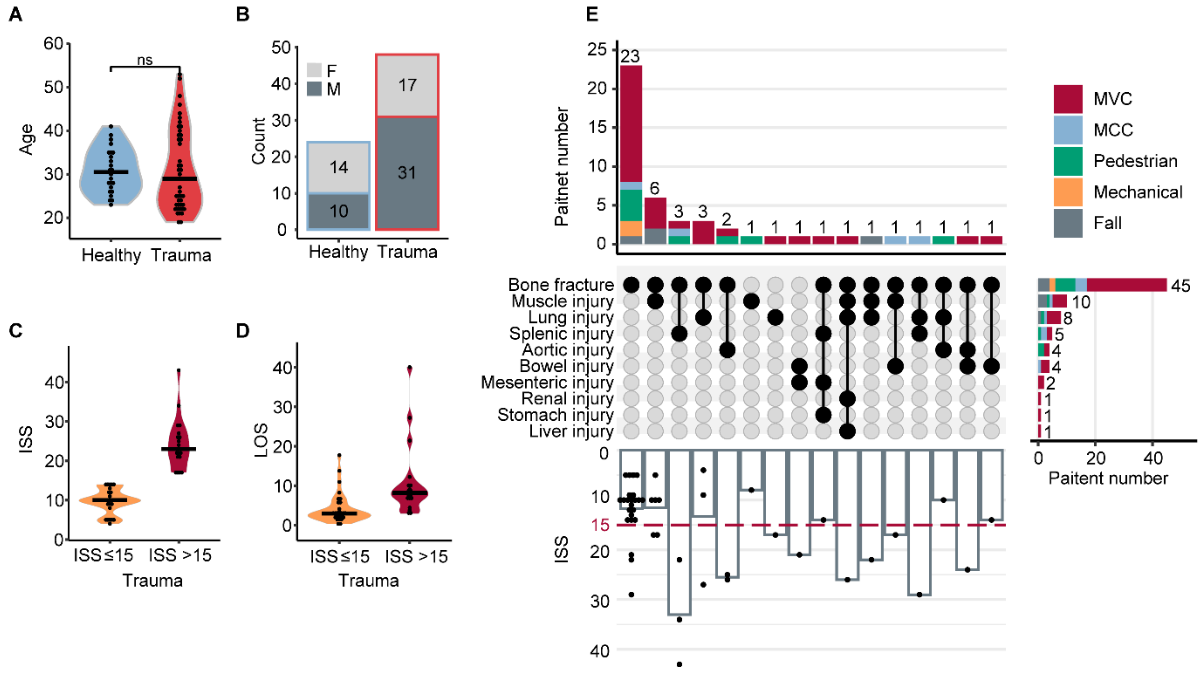


**Figure S1. Demographic and clinical information of the control and trauma cohorts, related to Table 1.**

**A.** Age distribution.

**B.** Sex distribution.

**C.** Injury Severity Score (ISS) distribution in the trauma cohort.

**D.** Length of Hospital Stay (LOS) in the trauma cohort.

**E.** Mechanisms of injury in trauma patients and their corresponding ISS score. The top bar plot graph shows the number of patients injured by various mechanisms: motor vehicle collision (MVC), motorcycle collision (MCC), pedestrian, mechanical force, and fall. The middle dot plot indicates injuries of specific body regions. The bar plot on the right shows the numbers of patients in each specific injury types. The bottom bar graph shows the ISS in patients with specific combination of injuries.

Unpaired student's t-test was used for significance test, and ns (non-significant) indicates p-value >0.05; ****, p-value<0.0001; **, p-value<0.01. Healthy: n=24; Trauma: n=48, ISS≤15: n=30, ISS>15: n=18.Bar indicates the median value.

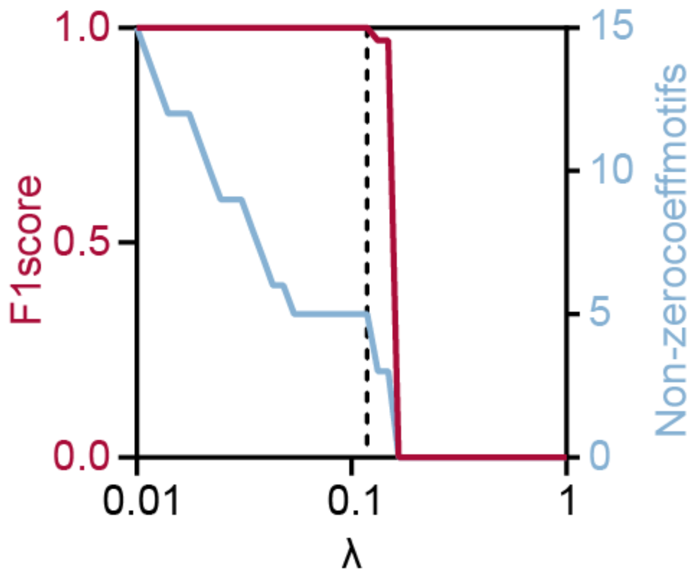


**Figure S2. Selection of essential motifs in LASSO model, related to Figure 2.**

The red curve represents the model accuracy (F1 score) while the blue curve represents the number of motifs in the model. The dashed vertical line indicates the optimal hyperparameter (λ) for the LASSO model, with which the model achieved the highest F1 score with the fewest motifs needed.

**A. Test data set 1 (20 mouse miRNAs)**

**Tested pro-inflammatory (13)**

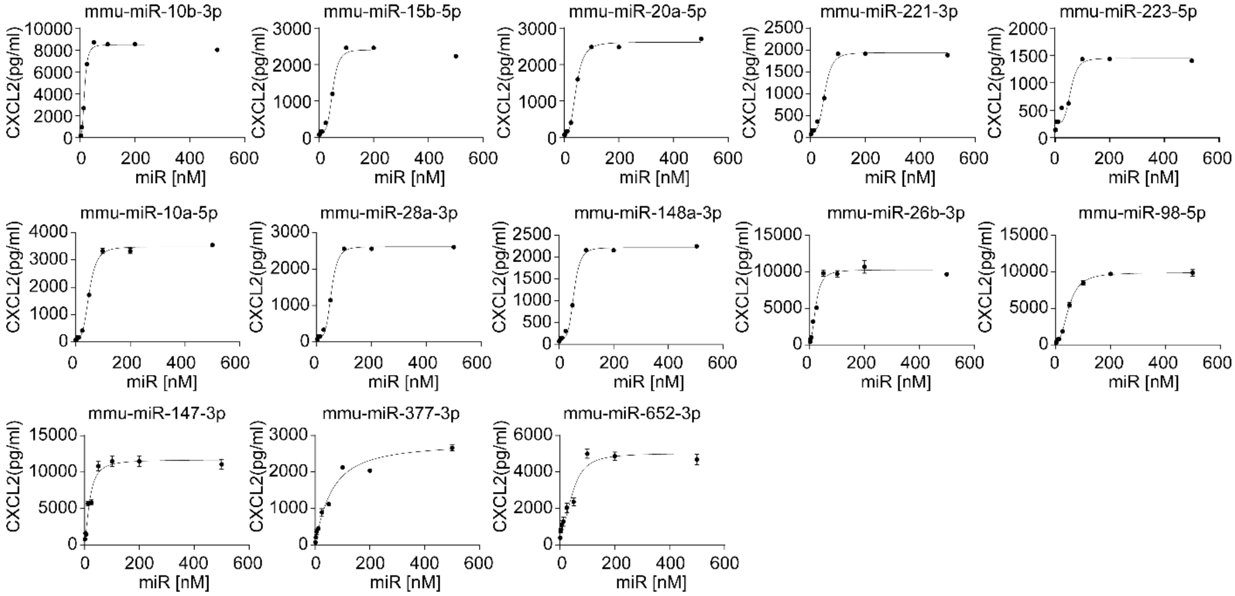


**Tested non-inflammatory (7)**

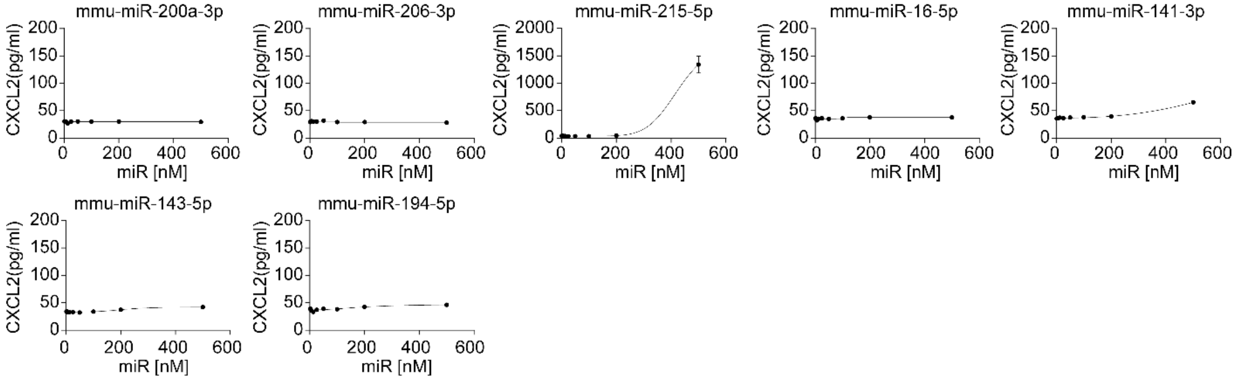


*(continued)*

### B. Test data set 2 (38 human miRNAs)

#### Tested pro-inflammatory (25)

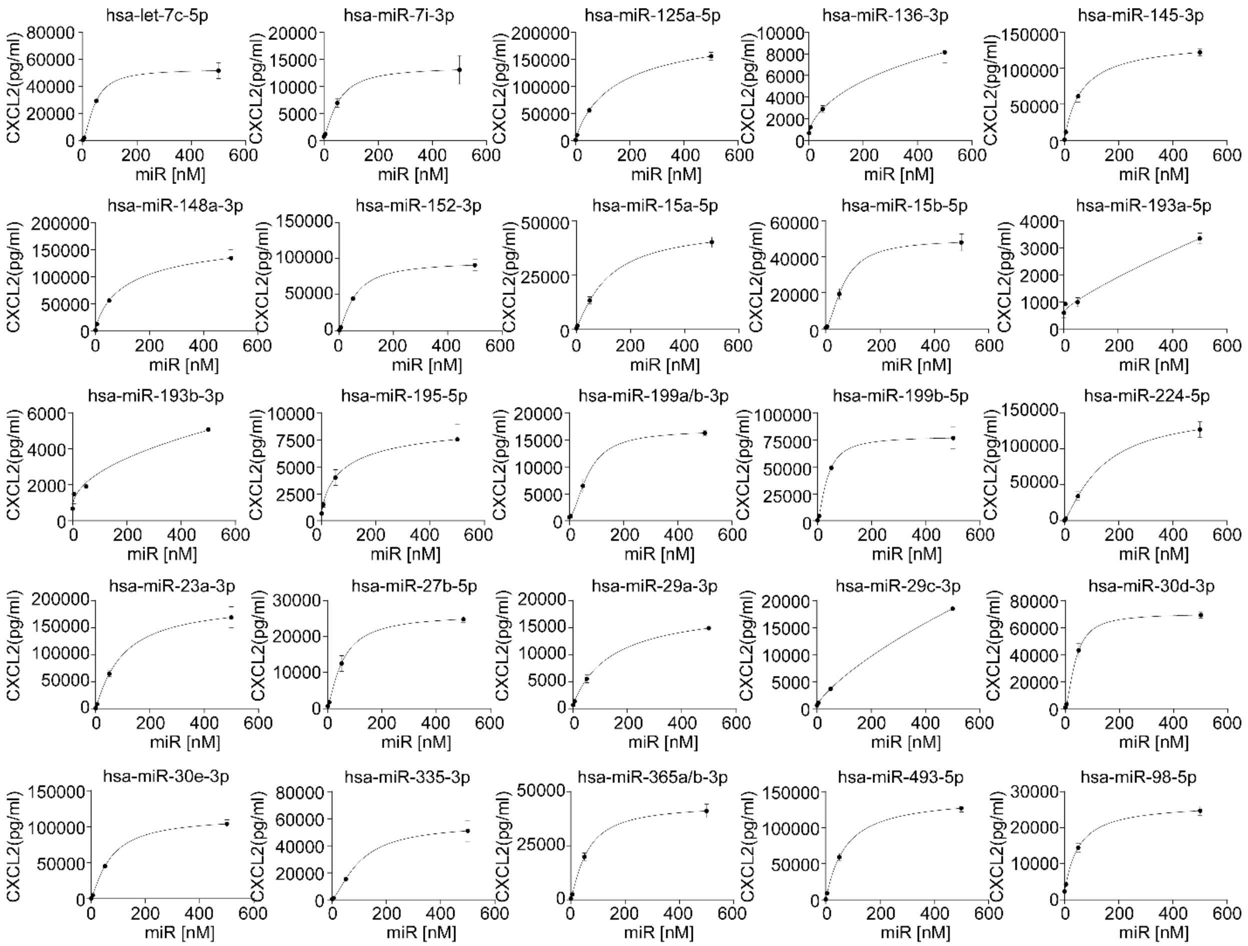


#### Tested non-inflammatory (13)

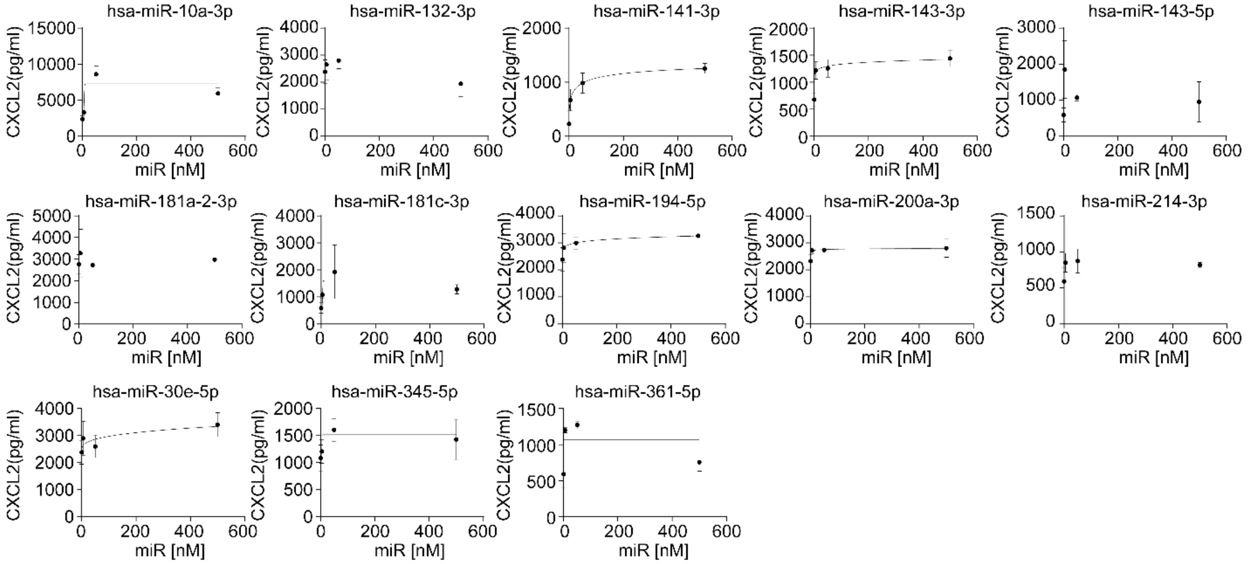

**Figure S3. Testing of miRNA mimics for CXCL2 production in cultured macrophages, related to Figure 2.**

A. Mouse MBDMs were treated with mouse miRNA mimics from the "test data set 1". Eighteen hours later, culture media were assayed for CXCL2 production using ELISA.
B. Mouse Raw 264.7 macrophage cultures were treated with human miRNA mimics from the "test data set 2".

Fitted non-linear regression with four parameters was shown as solid curves.

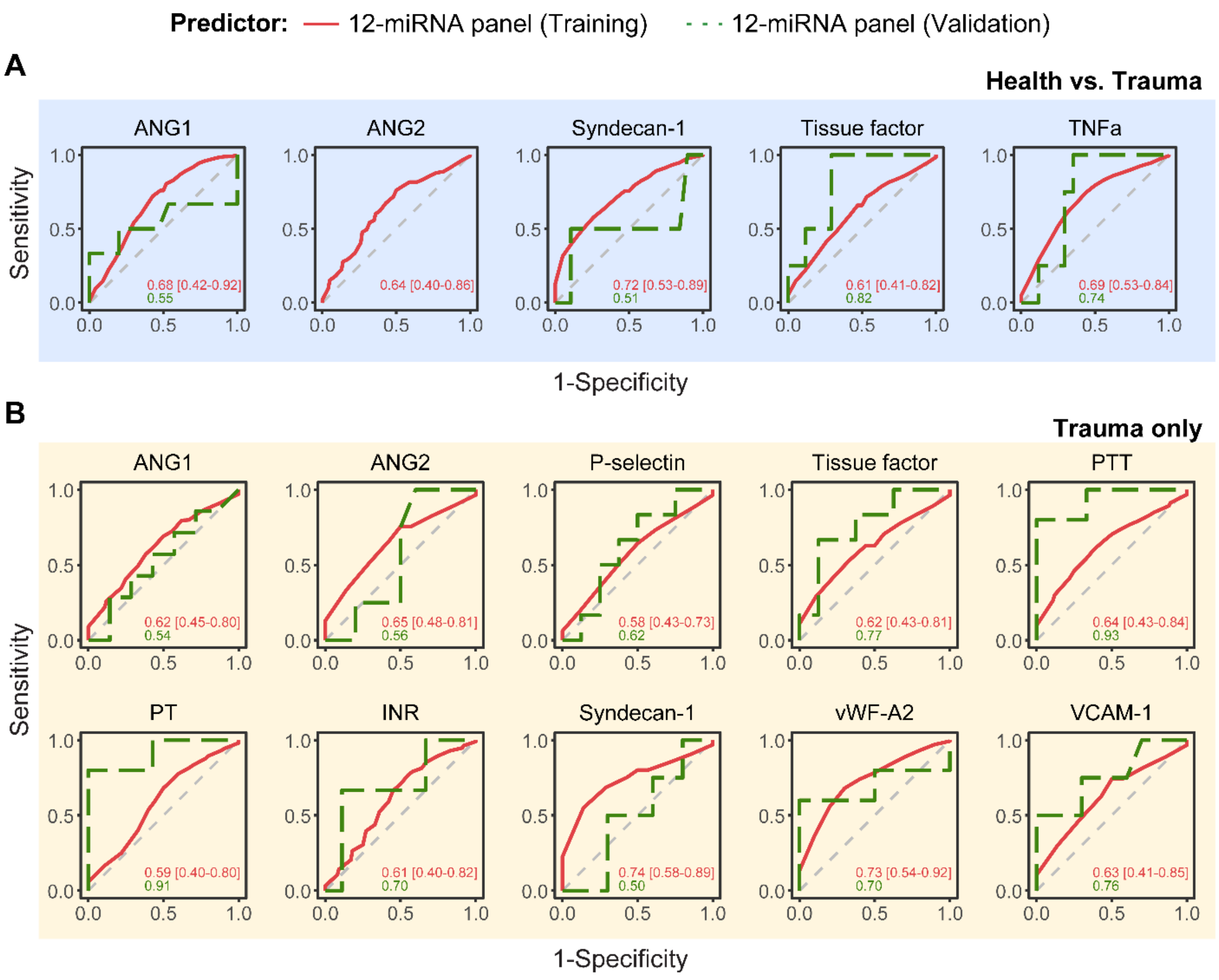


**Figure S4. ROC analysis for injury prediction, related to Figure 6.**

A. ROC curves for injury markers that remain normal or were slightly elevated in the trauma cohort.

B. ROC curves for identifying severe injury.

Training set (trauma, n=34; control, n=17) and 12-miRNA panel in Validation set (trauma, n=14; control, n=7) are shown. Random Forest regression was used for modeling within the Training set, with 100 times 2-fold cross-validation. AUROC values were presented with 95% confidence intervals.

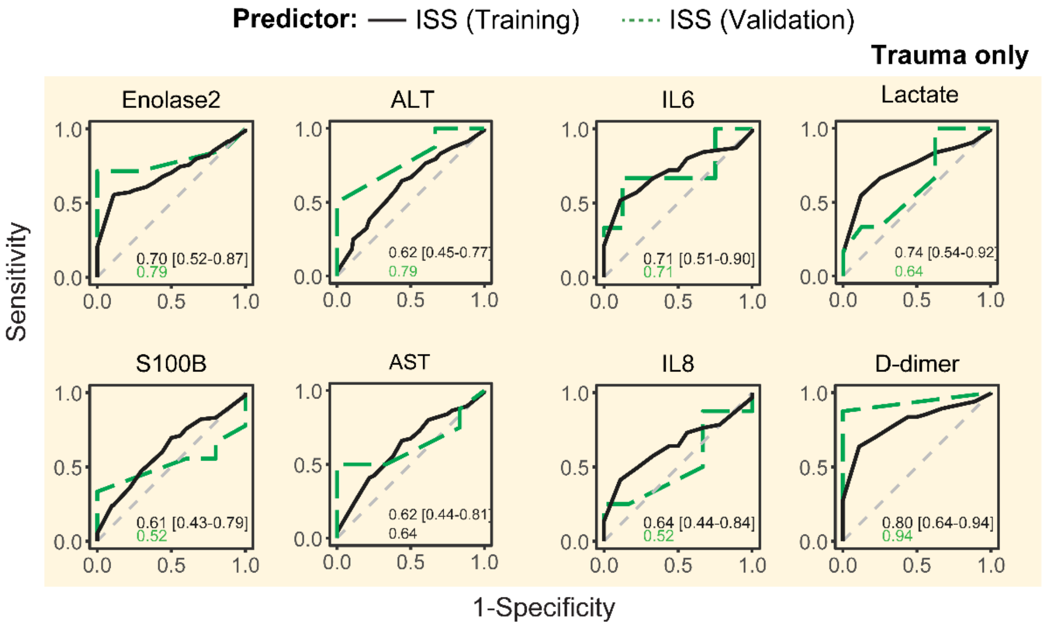


**Figure S5. Prediction of injury severity by ISS, within trauma cohorts, related to Figure 8.**

ROC curves of injury severity score (ISS) as predictor to severity of indicated injury markers (divided by the median level). Training set (n=34), validation set (n=14)

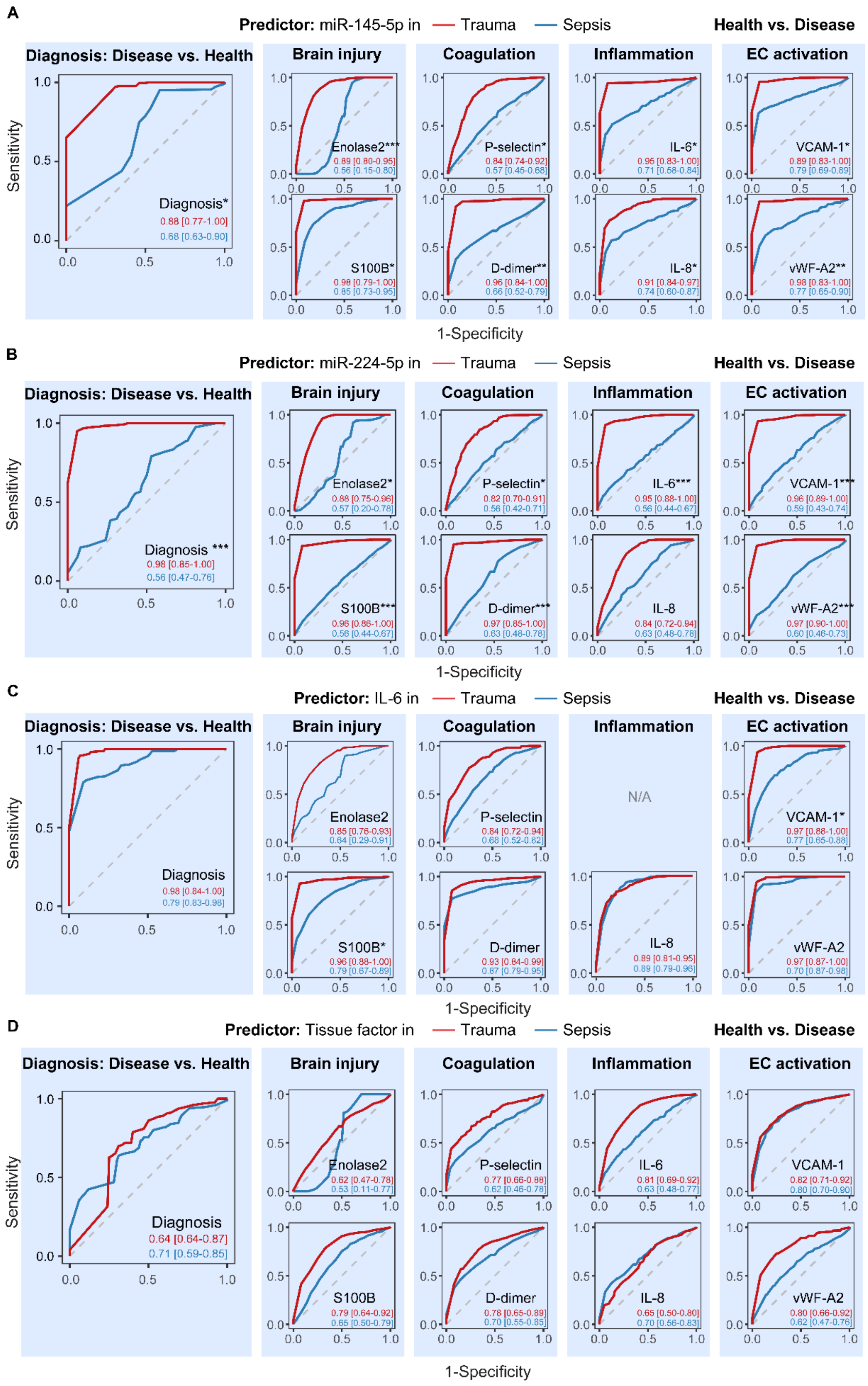

A
Predictor: miR-145-5p in — Trauma — Sepsis
Health vs. Disease
Diagnosis: Disease vs. Health
Sensitivity
Diagnosis*
0.88 [0.77-1.00]
0.68 [0.63-0.90]
Brain injury
Coagulation
Inflammation
EC activation
Enolase2***
0.89 [0.80-0.95]
0.56 [0.15-0.80]
P-selectin*
0.84 [0.74-0.92]
0.57 [0.45-0.68]
IL-6*
0.95 [0.83-1.00]
0.71 [0.58-0.84]
VCAM-1*
0.89 [0.83-1.00]
0.79 [0.69-0.89]
S100B*
0.98 [0.79-1.00]
0.85 [0.73-0.95]
D-dimer**
0.96 [0.84-1.00]
0.66 [0.52-0.79]
IL-8*
0.91 [0.84-0.97]
0.74 [0.60-0.87]
vWF-A2**
0.98 [0.83-1.00]
0.77 [0.65-0.90]
1-Specificity
B
Predictor: miR-224-5p in — Trauma — Sepsis
Health vs. Disease
Diagnosis: Disease vs. Health
Sensitivity
Diagnosis ***
0.98 [0.85-1.00]
0.56 [0.47-0.76]
Brain injury
Coagulation
Inflammation
EC activation
Enolase2*
0.88 [0.75-0.96]
0.57 [0.20-0.78]
P-selectin*
0.82 [0.70-0.91]
0.56 [0.42-0.71]
IL-6***
0.95 [0.88-1.00]
0.56 [0.44-0.67]
VCAM-1***
0.96 [0.89-1.00]
0.59 [0.43-0.74]
S100B***
0.96 [0.86-1.00]
0.56 [0.44-0.67]
D-dimer***
0.97 [0.85-1.00]
0.63 [0.48-0.78]
IL-8
0.84 [0.72-0.94]
0.63 [0.48-0.78]
vWF-A2***
0.97 [0.90-1.00]
0.60 [0.46-0.73]
1-Specificity
C
Predictor: IL-6 in — Trauma — Sepsis
Health vs. Disease
Diagnosis: Disease vs. Health
Sensitivity
Diagnosis
0.98 [0.84-1.00]
0.79 [0.83-0.98]
Brain injury
Coagulation
Inflammation
EC activation
Enolase2
0.85 [0.76-0.93]
0.64 [0.29-0.91]
P-selectin
0.84 [0.72-0.94]
0.68 [0.52-0.82]
N/A
VCAM-1*
0.97 [0.88-1.00]
0.77 [0.65-0.88]
S100B*
0.96 [0.88-1.00]
0.79 [0.67-0.89]
D-dimer
0.93 [0.84-0.99]
0.87 [0.79-0.95]
IL-8
0.89 [0.81-0.95]
0.89 [0.79-0.96]
vWF-A2
0.97 [0.87-1.00]
0.70 [0.87-0.98]
1-Specificity
D
Predictor: Tissue factor in — Trauma — Sepsis
Health vs. Disease
Diagnosis: Disease vs. Health
Sensitivity
Diagnosis
0.64 [0.64-0.87]
0.71 [0.59-0.85]
Brain injury
Coagulation
Inflammation
EC activation
Enolase2
0.62 [0.47-0.78]
0.53 [0.11-0.77]
P-selectin
0.77 [0.66-0.88]
0.62 [0.46-0.78]
IL-6
0.81 [0.69-0.92]
0.63 [0.48-0.77]
VCAM-1
0.82 [0.71-0.92]
0.80 [0.70-0.90]
S100B
0.79 [0.64-0.92]
0.65 [0.50-0.79]
D-dimer
0.78 [0.65-0.89]
0.70 [0.55-0.85]
IL-8
0.65 [0.50-0.80]
0.70 [0.56-0.83]
vWF-A2
0.80 [0.66-0.92]
0.62 [0.47-0.76]
1-Specificity

**Figure S6. Single predictor model for diagnosis and organ injury prediction, related to Figure 8.**

ROC curves for disease diagnosis and abnormal level of organ injury markers by **A.** miR-145-5p, **B.** miR-224-5p and **C.** IL-6 and **D.** Tissue factor in trauma cohorts (red line, n=72, 48 trauma, 24 health) and sepsis cohorts (blue line, n=71, 47 sepsis, 24 health) using random forest modeling with cross validation.

**Table S1. Computer exhaustive search algorithm, related to Figure 2.**

**Motif algorithm: Sequence processing and *k*-mer generation**

**Input:**

- A list of sequences of 33 miRNAs (Training data set).
- Motif length, *k*, an integer, can be ranging from 2 to 10 in the miRNA sequence list.

**Output:** A set of 513,599 motifs with wildcards.

1. Convert each sequence to lowercase.
2. Define a prefix '!' and construct prefixed sequences. Then concatenate all the prefixed sequences into a single long string.

***Function***: *k-mer Generation Optimization Process*

**Input:** Motif length *k* (*k*-mer)

**Output:** A set of *k*-mers with wildcard substitutions.

3. Extract all possible substrings of length *k* (*k*-mers) from the concatenated sequence from step 2.
   - 3.a Consider the concatenated sequence as a single continuous string of length *L*.
   - 3.b For each position *i* in the concatenated sequence, where $0 \le i \le L-k$, extract a substring of length k starting from position *i* until all possible starting positions for a *k*-mer in the concatenated sequence have been considered, resulting in a total of $L$-$k$+1 *k*-mers.
   - 3.c Filter out *k*-mers containing the prefix character and duplicated *k*-mers.
4. Generate *k*-mers with wildcards.
   - 4.a Initialize a variable, *output_variable*=set(), to store *k*-mers with wildcard substitutions.
   - 4.b For each *k*-mer remained after step 3.c: Generate *k*-mers with wildcards at 1 to *k* positions. Each *k*-mer can have multiple wildcards, with a maximum of *k* wildcards per sequence. For each subset of positions, replace one or more characters with a wildcard (.).
   - 4.c Add each modified *k*-mer to *output_variable*. The use of Python's set structure ensures uniqueness and avoid redundant calculation.

**End Algorithm**

**Table S2. Pro-inflammatory motifs identified with a relaxed LASSO algorithm, related to Figure 2.**

| # | Motif | # | Motif | # | Motif | # | Motif | # | Motif |
|---|---|---|---|---|---|---|---|---|---|
| 1 | .UUC | 11 | A.U...U..G | 21 | GG.....U | 31 | U.GU...U.. | 41 | UUU. |
| 2 | U..UU. | 12 | AG.UU | 22 | GU....UU.. | 32 | U.GUG..... | | |
| 3 | .A.....UU. | 13 | AUU.....U | 23 | GU.UU | 33 | ....U.UC | | |
| 4 | G.UU. | 14 | AUUC | 24 | U.....GG | 34 | .UC...C.. | | |
| 5 | UU...U | 15 | .AUUC | 25 | U...C....U | 35 | UC..U | | |
| 6 | A......UC | 16 | AUUC.. | 26 | U...UC | 36 | UGGU | | |
| 7 | A......UUG | 17 | .AUUC.. | 27 | U...UC.... | 37 | UU...U.... | | |
| 8 | A...UUC | 18 | .G.....C.U | 28 | U..G.U. | 38 | UU.G | | |
| 9 | ..A..UU.. | 19 | G....U...U | 29 | .U..U.C..U | 39 | .....UUC | | |
| 10 | A.U...U... | 20 | G.UUG | 30 | U.GGU | 40 | ..UUC.... | | |
